# High remission rates and visual network normalization in severely traumatized children following medication-free, intensive inpatient psychotherapy

**DOI:** 10.64898/2026.03.21.26348507

**Authors:** Ludwig Ebeling, Maxim Korman, Julia Quehenberger, Catherina Dehmel, Verena Wagner, Stephan Goerigk, Michel Menzel, Liwen Yang, Anne Budke, Lukas Oberschneider, Julia Gollhammer, Sophia Stoecklein, Frank Padberg, Birgit Ertl-Wagner, Karl Heinz Brisch, Daniel Keeser

**Author notes:** These authors contributed equally to this work: Ludwig Ebeling, Maxim Korman, Birgit Ertl-Wagner, Karl Heinz Brisch, Daniel Keeser. In 2019, the Pediatric Psychosomatic Medicine and Psychotherapy, Dr. von Hauner Children’s Hospital, was reassigned to the Department for Child and Adolescent Psychiatry, Psychosomatics and Psychotherapy within the LMU University Hospital, Ludwig-Maximilians-University LMU; Munich, Germany.

## Abstract

Children exposed to severe childhood maltreatment often develop complex mental health disorders where standard treatments show limited efficacy. Current residential approaches combine psychopharmacological with behavioural interventions, yet the feasibility and clinical-neurobiological outcomes of intensive, medication-free psychotherapy have not been investigated in this population.

Our prospective study followed severely traumatized children (aged 6-13 years) with documented histories of changes and failures in placement.They completed an intensive 6-8 months inpatient treatment program (5 individual psychotherapy and 3 group therapy sessions per week with high caregiver-patient ratio) grounded in a novel, multimodal, attachment-based therapeutic framework. Medication was discontinued prior to treatment. The intervention group was compared to healthy controls and waitlist controls receiving treatment as usual.

Participants in the intervention group achieved high remission rates for dysregulated behaviour (Child Behaviour Checklist (CBCL) >60% post treatment, 50% on follow-up) and trauma-related symptoms (Parent Report of Post-traumatic Stress Symptoms (PROPS) >65% post treatment, >60% on follow-up). Within-group effect sizes for Total Problems Score, Externalising behaviour (both CBCL), Hyperactivity (Strengths and Difficulties Questionnaire) and trauma symptoms (PROPS) each exceeded Cohen’s d = 1.0 and were maintained at 6-month follow-up. Resting-state fMRI identified significant functional reorganization in visual processing networks. Atypical correlation patterns between visual network activity and symptom severity resolved following treatment, yielding patterns comparable to those of healthy controls. These pilot findings provide initial evidence of the feasibility and effectiveness of intensive, medication-free, attachment-based inpatient treatment to promote clinical remission and neurobiological normalization in severely traumatized children.

## 2. Introduction

Childhood maltreatment (CM) is widely recognized as one of the most significant preventable risk factors for adverse health outcomes across the lifespan^1–4^, imposing substantial societal costs that reach up to 6% of GDP in European countries^5^. The relationship between CM and later health outcomes follows a robust dose-response pattern: individuals exposed to four or more types of maltreatment face strongly elevated risks for adverse health outcomes compared to those experiencing a single type of CM^6,7^. A similar pattern is observed in neurobiological findings, where severity of exposure tends to be reflected in its impact on brain function and structure^8^.

The effects of cumulative CM burden manifest across multiple life domains and generations. The effects of cumulative CM burden manifest across multiple life domains and generations. For example, high childhood adversity exposure is associated with increased juvenile violent behaviour^9^, and parental adversity has been shown to predict behavioural problems in offspring through its effects on parenting style^10^ - suggesting an intergenerational cycle in which adversity shapes parenting, which in turn shapes the next generation’s health and behaviour. Crucially, childhood and adolescence represent a window of opportunity during which early adverse influences may be mitigated before secondary negative outcomes ensue, particularly when interventions are designed to harness the plasticity of the developing brain. Investment in early interventions therefore promises “a triple dividend, with benefits for young people today, the adults they will become, and the next generation of children they will parent“^11^.

While the benefits of focusing on a highly burdened population are evident, the challenges are substantial. While outpatient treatments for PTSD such as Trauma-Focused Cognitive-Behavioural Therapy (TF-CBT)^12–15^ and Eye-Movement Desensitization and Reprocessing (EMDR)^16–18^ in children or Cognitive Processing Therapy (CPT) among adolescents^19^ are well established, evidence for treatment effects regarding complex traumatization are sparse^20^. Individuals with a complex trauma history require significant modifications to standard protocols^21^, addressing substantial internalizing and externalizing problems, often manifested in pervasive difficulties in emotion regulation and interpersonal functioning. Here, treatments that include caretaker involvement consistently yield superior effect sizes^22^.

The more severe the emotional and behavioural difficulties in children are, the less likely they are to remain in stable settings - be it parental homes, foster care, or schools - resulting in repeated experience of disrupted attachment and rejection. As a consequence of this escalating instability, residential treatment centers (RTC) are often the last resort. However, RTC settings and treatments differ so widely they have been likened to a “black box”^23^. The vast majority of youth in residential treatment have a history of multiple traumatic events, which is associated with a broad variety of functional impairments^24^. This history itself has been proposed as a marker that a more intensive intervention is required^24^. While several RTCs have employed trauma-informed care interventions in recent years, the empirical literature remains limited. Notably, the vast majority of published intervention approaches originate from the United States, only one study to date has included a European site^25^.

Standard treatment in this population relies heavily on psychotropic medication - despite concerns regarding limited effectiveness in complex trauma, potential side effects, and long-term effects on development^26,27^ - while the clinical potential and neurobiological mechanisms of intensive psychotherapy without pharmacology remains largely unexplored. Pharmacotherapy can be a very effective tool to reduce severe symptoms, but its potential physiological and neurobiological side effects should not be underestimated^26–28^.

Further, time-out placements (TOP) remain widely established in inpatient treatments as a common short-term intervention. These placements are intended to address acute affect dysregulation and to temporarily separate children exhibiting severe behaviours from the environment in which these behaviours occur. However, they have been shown to have detrimental effects on subsequent behaviour and overall placement stability within youth care facilities^29^.

Interventions differ in both their clinical and developmental consequences, as well as the neurobiological imprints they impose on the developing brain. This highlights the need for neuroimaging-based investigations to determine whether and how these interventions alter functional brain organization. To the best of our knowledge, no published study has yet examined the effects of an intensive inpatient treatment through a longitudinal neuroimaging lens. While the latter holds specific challenges in young patients^30^, it can potentially deliver objectifiable biomarkers to monitor treatment effects and deepen our understanding of healthy and traumatized development trajectories. From a neurodevelopmental perspective, some of the published literature aligns with models of maltreatment-related brain adaptation. Childhood maltreatment shapes brain development in an experience-dependent manner, with particular effects on sensory and perceptual systems involved in threat detection^31^. Alterations in primary and secondary visual spots (e.g., V1 and V2) could be hypothesised as adaptive responses that may facilitate heightened vigilance in adverse environments rather than as fixed deficits^31–33^. Against this background, visual processing networks may constitute a foundational neurobiological substrate for trauma-related adaptations and a particularly sensitive target for therapeutic change.

To address these gaps, the present study implemented a time-intensive and attachment-based trauma-informed approach at a specialized inpatient trauma unit. We evaluated treatment effects through a broad battery of clinical tests and longitudinal resting-state functional connectivity (rs-FC). Importantly, this study implemented an exclusively non-pharmacological treatment approach, allowing the psychotherapeutic effects on the developing brain to be assessed without the confounding influence of psychotropic medication. Our approach implements a range of core treatment outcome mediators validated in a recent systematic review^34^, such as patient engagement, caregiver support, therapeutic alliance and collegial collaboration. We hypothesized that an intensive, multimodal trauma-informed treatment approach (1) reduces behavioural and affective self-regulation difficulties and (2) drives a measurable reorganization of functional brain networks, aligning them more closely with those observed in healthy controls.

## 3. Methods

### Study design

We conducted a controlled clinical trial, testing a high-intensity psychotherapeutic intervention for severely traumatized children at the Department of Pediatric Psychosomatic Medicine and Psychotherapy, Dr. von Hauner Children’s Hospital, Munich University Hospital, Ludwig-Maximilians-University (LMU), Munich Germany. The study received approval from the Ethics Committee of the LMU (Approval Number: 326-12).

The study used a quasi-experimental design comprising an intervention group, a wait list control group, and a healthy comparison group. The intervention group received intensive psychotherapy, with 5 sessions of individual therapy weekly, lasting 6-8 months on average and consisting of equal parts of psychodynamic and trauma therapy.

Assessments for the intervention group were completed at baseline (T1), post treatment (T2, 6-8 months), and at follow-up (T3, 12-14 months after baseline). The healthy control group was matched to patients on age and gender and assessed at baseline (T1) and post treatment (T2). See the work flow diagram in Figure 1.

**Figure 1:**
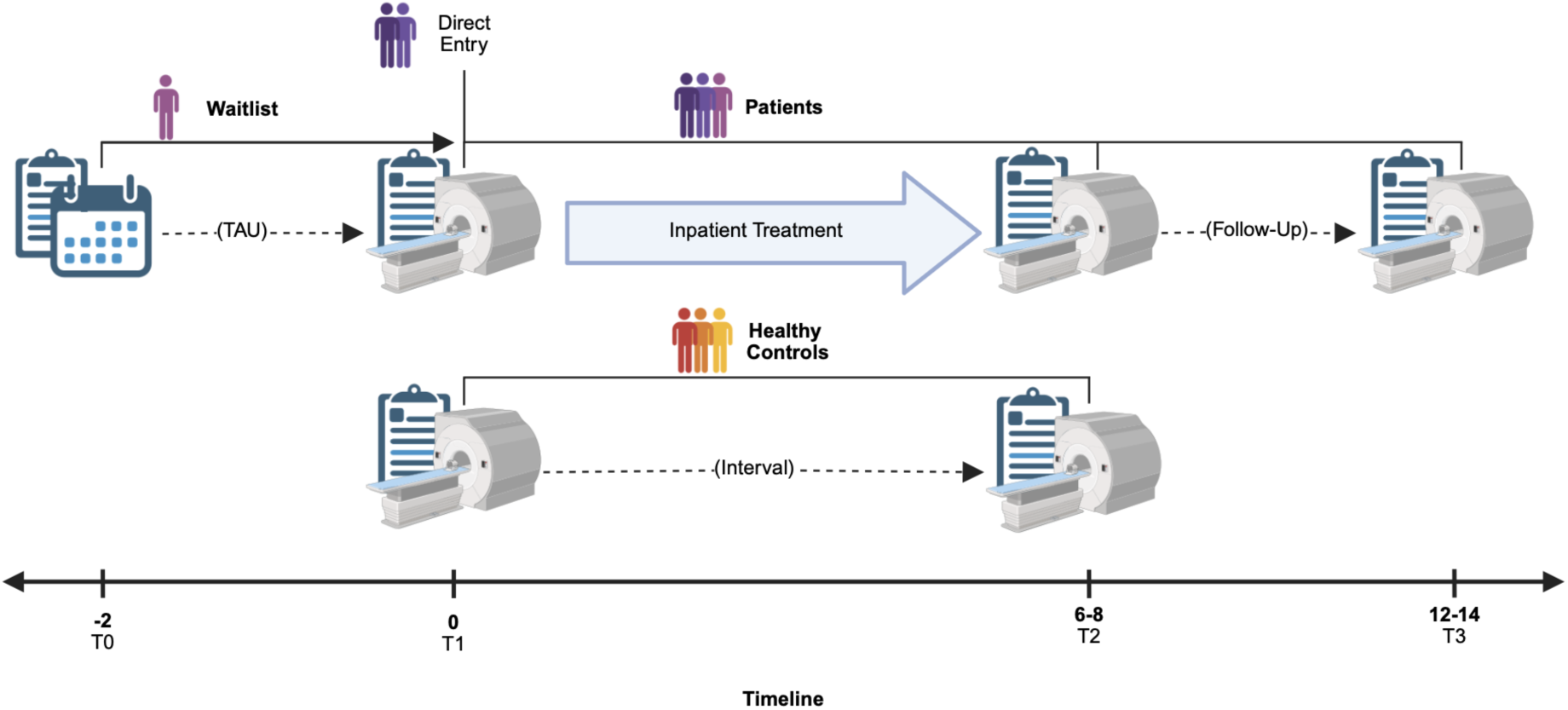
Study workflow diagram. The timeline (horizontal axis) illustrates the study phases relative to the start of the active intervention (T1, month 0). Waitlist & treatment as usual (TAU): A subset of patients enters the study at T0 (month -2). These participants serve as a within-subject control, receiving standard outpatient care (TAU) for at least two months to establish a baseline for symptom stability prior to the active intervention. Inpatient Treatment (intervention): At T1 (baseline), all patients - including those from the waitlist and those entering via Direct Entry - undergo baseline MRI scanning and clinical assessment. This marks the beginning of the intensive, multimodal inpatient psychotherapy program (approx. 6–8 months, 5 hours individual psychotherapy and 3 hours group therapy per week). Post-Treatment & Follow-Up: Patients undergo a second scan at T2 (months 6–8) to evaluate treatment effects and a final follow-up scan at T3 (months 12–14) to assess the maintenance of clinical and neurobiological changes. *Note: Patient cohort is represented in purple and healthy controls in orange. Dashed arrows symbolise treatment as usual (TAU), Intervals, or Follow-Ups without active study intervention*.

Additionally, a subgroup of the patients in the intervention group were assigned to the waitlist first due to constraints on resources and were assessed at least two months prior to starting treatment (T0). During this time and until switching to the intervention group, they received treatment as usual (TAU). TAU typically comprises outpatient psychotherapy (maximum of two sessions per week) combined with pharmacotherapy.

### Participants

Children aged 6-13 years were recruited from public child welfare institutions between (2012-2019). Contact with orphanages, caregiving agencies, and youth welfare offices were established to present our study. Additionally, patients presenting to our outpatient clinic were screened for inclusion criteria.

Participants underwent comprehensive clinical interviewing with a child psychiatrist (see Table S1 for detailed clinical assessment). Assignment to the intervention group occurred for individuals displaying severe symptoms alongside documented evidence of childhood maltreatment or considerable adversity assessed in initial phone screening, via questionnaires (Lifetime Incidence of Traumatic Events, Medical History Questionnaire) and through structured clinical interviews. While presenting symptoms were diverse, a unifying feature was recurrent placement disruptions across multiple settings (including foster care and schools) driven by severe behavioural challenges. The most common types of adversity were hospitalized family members (61%), separation from family (42.9%), death of a family member (38.1%), physical abuse (33.3%), and divorce of parents (28.6%). The wait-list control group consisted of participants who met the same criteria as the intervention group but were temporarily placed on a waiting list before receiving the intervention.

Exclusion criteria for the intervention group included: Acute suicidality or behaviour that posed a danger to others, substance use disorders, profound developmental disorders with intellectual disability (IQ < 85), autism spectrum disorders, somatic conditions including epilepsy and metabolic disorders, MRI-related contraindications such as the presence of a pacemaker, implanted defibrillator, other ferromagnetic implants, or claustrophobia, and insufficient or no proficiency in the German language.

The healthy control group was recruited through flyers and posters and underwent the same clinical assessment procedures as the patients. Additional exclusion criteria for the healthy control group included the presence of any psychiatric diagnosis according to ICD-10^35^ and any history of outpatient psychiatric or psychotherapeutic treatment. Written informed consent was obtained from parents or legal guardians, and age-appropriate assent was obtained from all child participants.

### Intervention

Patients in the intervention group received intensive inpatient psychotherapeutic treatment embedded within a trauma-informed and attachment based framework for 6-8 months in a dedicated treatment unit at the Dr. von Hauner Children’s Hospital of the University Munich^36^. Table 1 provides an overview of our intensive treatment program compared to treatment as usual. The multimodal treatment comprised the following core components:

**Table 1:** Comparison of Core Clinical and Structural Features Between Treatment as usual and the Intensive Inpatient Treatment Unit. . The table contrasts standard inpatient psychiatric care with the specialized attachment-based, trauma-informed intervention across key therapeutic, pedagogical, and organizational domains. *Note: **PDS**, Posttraumatic Diagnostic Scale; **IES**, Impact of Event Scale; **SCL-90,** Symptom-Checklist-90; **BDI**, Beck-Depressions-Inventar; **TAQ**, Traumatic Antecedents Questionnaire*.

| Conceptual feature | Treatment as usual<br>(inpatient unit) | Intensive inpatient treatment unit |
| --- | --- | --- |
| **Attachment** | Case-handling by lead therapist or physician. | Designated and consistent primary therapist, caregiver, and physician as attachment figures. At admission, each subteam also designates a backup staff member and discusses this with the patient to cover both scheduled and rarely unforeseen absences of team personnel. |
| **Treatment** | Symptoms- and resource-oriented, stabilisation, behavioural regulation, crisis management. Predominantly conducted through group formats employing fixed treatment protocols. | Based on psychodynamics and attachment theory. Trauma-informed intervention. Personalised to match individual symptom profiles and trauma backgrounds. Prioritize sustained engagement with the children to facilitate co-regulatory processes throughout episodes of intense emotional dysregulation (triggered attachment responses). |
| **Pedagogical focus** | Structure, rules, boundaries. | Strengthening of patients' capacities for healthy attachment in a stable and secure environment. Fostering understanding across the entire eco-system (patient, family, treatment staff). |
| **Crisis intervention** | Time-out, withdrawal, medication. | Time-intensive attachment regulation (1:1 or 2:1 support), emergency psychotherapy sessions. |
| **Team reflection sessions** | During daily shift handovers. Ideally conducted weekly case supervision with limited time per patient or selected single cases. | Very frequent: daily case conferences (1.5hrs), plus twice-weekly case supervision (2hrs) and once a week team supervision (2hrs) to regulate and understand stress-related team-processes. Occasionally: extended trauma-informed case supervision over several days. |
| **Staffing** | Larger groups, standard ratios. | Small groups (appr. 5-6 children, 3 staff); minimum 2:1 ratio. |
| **Patient participation** | Limited. | Daily patient conferences with focus on active child participation. |
| **Transference/countertransference** | Rarely discussed. | Central to self-reflection of staff, therapeutic understanding and action, intends to reduce risk of potential retraumatization. |
| **Schooling** | External schooling, no schooling or a clinical school for all inpatients spanning all clinic departments. | Schooling took place in a special classroom only for the patients of the treatment group, often in 1:1 or 1:2 settings. In a situation of acting out, children were co-regulated by teachers as well as by staff. Children could return to the ward for further co-regulation if the co-regulation in the classroom was not successful. |
|  |  | Close supervision of teachers several times per week with the perspective to restore participation in public schools post treatment. Teachers were considered to be important “members of the treatment team”. |
| **Interdisciplinary collaboration** | As needed. | Daily integration across therapy, education, physicians and nursing. |
| **Pharmacological treatment** | Often used. | Not used. |
| **Transgenerational symptoms** | Examined as a supplementary consideration during parent consultation. | Comprehensive assessments of caregivers mental and health and trauma history (e.g. PDS, IES, SCL-90, BDI, TAQ). If needed, caregivers were referred to individual outpatient therapy. |

#### Individual psychotherapy

Participants received at least four hours of individual psychotherapy weekly (mean = 5 hours), equally divided between psychodynamic and trauma-focused approaches. Treatment was delivered by psychotherapists, psychotherapists-in-training and trauma-trained physicians. Emergency sessions were provided upon request by the patient or when clinically indicated by the patient’s primary psychotherapist or physician.

#### Milieu therapy

Continuous therapeutic milieu was maintained through a primary caregiver system staffed by nurses, social pedagogues, nursing students, and volunteers. A staff-to-patient ratio of at least 1:2 was maintained at all times. It was essential to maintain the therapeutic space even in moments of crisis:

“Time-intensive” interventions replaced traditional time-out procedures, providing 1:1 or 2:1 co-regulation support during acute dysregulation episodes, with emergency sessions available from key therapists as needed.

#### Adjunctive therapies

Patients chose a creative therapy - such as music, art, movement, or sports therapy - as supplementary individual therapy and participated weekly in all remaining sessions in group format. Additional interventions included an observation-based mentalizing intervention (B.A.S.E.-Babywatching^37^) and Respectare applications (structured, child-controlled touch interventions such as hand or back massage) to foster safe proximity and positive bodily experiences.

#### Family involvement

Parents attended weekly counseling sessions with their child’s two primary therapists and participated in progress evaluation sessions with the medical director every six weeks. Weekly home leave (Saturday–Sunday) allowed for behavioural skills practice in natural settings and provided continuous real-life feedback regarding the therapeutic progress.

#### Staff support and development

The entire treatment team received case supervision twice per week in addition to weekly multidisciplinary team supervision. Each session lasted two hours. All supervision was conducted by experienced external supervisors. Daily clinical case conferences (1.5 hours) provided additional exchange with the medical director. Occasionally the whole team attended trauma-informed case supervision over several days by external experts. Continuous in-service training focused on recognizing and utilizing staff emotional responses to prevent retraumatization and maintain team resilience. Conceptual review meetings during which staff collaboratively proposed and discussed program refinements took place every six weeks.

#### Exclusion of pharmacological treatment

Psychopharmacological treatment was not used throughout the program; any pre-existing medication was appropriately discontinued prior to admission.

### Clinical assessments

Participants self-assessed their depressive symptoms using the Depression Inventory for Children and Adolescents (DIKJ^38^), all other clinical questionnaires were answered by parents and other primary caregivers. Caregivers were asked to fill out the Parent Report of Post-traumatic Symptoms (PROPS^39^), a 30-item questionnaire examining the presence and intensity of possibly trauma-related symptoms throughout last week. Furthermore, the Child Dissociative Checklist (CDC^40^) was used to screen for dissociative symptoms throughout the past 12 months and the Lifetime Incidence of Traumatic Events (LITE-P^41^) provided information on traumatic life experiences. The Strength and Difficulties Questionnaire (SDQ^42^) and the Child Behaviour Checklist (CBCL^43^) were completed by parents and other primary caregivers. Given the heterogeneous symptom presentation typical of early-traumatized patients, we utilized the total scores from these two measures as quantitative indicators of overall symptom severity. Additionally, the hyperactivity subscale of the SDQ and the internalizing and externalizing behaviour subscales of the CBCL provided more fine-grained characterization of symptom profiles.

Furthermore, the Screen for Child Anxiety Related Disorders (SCARED^44^) was administered. The Wechsler Intelligence Scale for Children–Fourth Edition (WISC-IV^45^) was used at baseline for the intervention group as part of the initial diagnostic evaluation. To mitigate learning effects, our assessments for the remaining measurements, as well as the healthy control group, specifically focused on evaluating the Working Memory Index (WMI) of the WISC-IV.

### Neuroimaging

Resting-state functional connectivity was analysed across large-scale brain networks using linear mixed-effects models. Potential confounding factors including gender, relative head motion, handedness, presence of support persons during scanning, and scanner type were included as covariates in the regression models, with their influence quantified using partial omega-squared effect sizes. P-values were adjusted using Hommel’s method to control for multiple comparisons.

#### MRI Acquisition

Imaging was conducted on 3T Siemens scanners (Magnetom Verio, Skyra) at the Institute for Clinical Radiology, Ludwig-Maximilians-University Munich, using 32-channel head coils. Full acquisition details are provided in Table S7. Scanner assignment was balanced across groups to minimize systematic bias (Table S2).The protocol included: T1-weighted 3D MPRAGE (voxel size 1.0×1.0×1.0 mm, TR=2100 ms, TE=4.37 ms, FA=7°, 160 slices, 4:18 min) and resting-state fMRI (voxel size 3.0×3.0×3.0 mm, TR=3000 ms, TE=30 ms, FA=80°, 46 slices, 200 volumes, 10:06 min). During resting-state acquisition, participants were instructed to keep eyes closed and avoid focused thought. Total scan time was approximately 40 minutes.

To minimize anxiety and motion artifacts, children were accompanied continuously by a primary caregiver and a psychotherapist or physician. Procedures included pre-scan familiarization with the scanner environment, child-appropriate educational materials, individualized hearing protection, and the option for a parent/caregiver to remain in the scanner room during acquisition. To ensure data viability in this highly dysregulated cohort, a known attachment figure (e.g. primary therapist) remained in the scanner room across all scanning sessions to provide co-regulation when needed, a protocol essential for avoiding sedation.

#### Preprocessing

Data was processed using FSL 6.0 (FMRIB Software Library). Preprocessing included: brain extraction (BET), removal of first four volumes for signal stabilization, slice-timing correction, motion correction (MCFLIRT, 6 degrees of freedom), spatial smoothing (5mm FWHM), intensity normalization, and high-pass temporal filtering (σ=50s). Images were co-registered to individual T1-weighted scans and normalized to MNI152 standard space (2mm isotropic) using FLIRT.

#### Motion Correction and Quality Control

Datasets with mean relative displacement (MRD) >0.55mm were excluded^46^. Single-subject independent component analysis (ICA) was performed using FSL MELODIC (20 components). Components were manually classified as signal or noise following established criteria^47^, and noise components were regressed out using FSL_regfilt. This approach improved temporal signal-to-noise ratio from 14.11 (raw) to 52.77 (post-processing), representing a 274% improvement - acceptable given the pediatric clinical population and expected motion artifacts.

#### Resting-State Network Analysis

Group ICA was performed using FSL MELODIC with probabilistic ICA and 20 dimensions, following optimization procedures to maximize network clarity while minimizing artifact-related lateralization. Networks were identified using the six-network taxonomy: occipital (visual), pericentral (somatomotor), dorsal frontoparietal (attention), lateral frontoparietal (control), midcingulo-insular (salience), and medial frontoparietal (default mode)^48^. A spatial overview of these identified resting-state functional networks is shown in Figure S1. Dual regression was applied to extract subject-specific spatial maps. Activated voxels were quantified using a threshold of z>4 with binary masking.

### Statistical analysis

Statistical analyses were performed using R software (Version 4.2.1)^49^. Linear mixed-effect models were conducted using the lme4 package^50^ and lmerTest^51^. For each outcome variable (clinical measures or neural network activation), we fitted a separate model, including a random intercept for each subject. Group was used as the between-subjects factor (patient vs. healthy control) and Measurement as the within-subjects factor (three timepoints). Both main effects and their interaction (grouping x measurement) were included as fixed factors. Potential confounders (age, gender and additionally for network activation: scanner type, presence of a supporting person during scan, handedness, motion plus interaction terms for grouping*scanner type and grouping*motion) were included as covariates. Models were estimated using REML. Statistical significance was evaluated at α = 0.05.

Partial omega-squared (ω²□) was selected to quantify the effect size for explained variance, based on its superior reliability with smaller sample sizes^52,53^. To address potential bias from small and unequal sample sizes, fixed-effect tests used Kenward–Roger approximation for degrees of freedom^54^. Post-hoc pairwise comparisons were performed using the R package emmeans^55^, p-values were adjusted using the Hommel method, which is more conservative than the FDR method while having improved disjunctive performance compared to Bonferroni, and maintains reliability with non-normal distributions^56^. All regression models were diagnostically evaluated for homoscedasticity and linearity through both graphical and statistical approaches. Variance inflation factors were calculated to identify problematic multicollinearity (threshold VIF > 10) among independent variables.

Additionally, we computed a meta-effect size (d_mean_) by aggregating individual Cohen’s d values across all clinical outcome variables (within-subject-factors) to provide a single summary effect. For exploratory correlation analyses, we employed Pearson’s correlation coefficient without p-value adjustment due to the small sample size and the hypothesis-generating nature of these analyses. Remission rates on clinical variables (Table S8) were calculated from complete-case samples for participants who exceeded clinical cutoff at baseline and subsequently dropped to subclinical levels at both post-treatment and follow-up assessments. BioRender (https://BioRender.com) was chosen to design the workflow diagram.

## 4. Results

### Cohort description

The overall sample included 39 participants, who were assessed at up to 4 timepoints. The intervention group initially consisted of 27 patients at baseline (T1) assessment, of whom 7 dropped out before post-treatment testing (T2). Of these, 3 dropped out because their families relocated, and 4 required transfer to secure psychiatric units due to symptom severity (two for externalizing behaviours with risk to others, two for acute suicidality). Given the severe clinical burden of our cohort, the proportion of participants requiring transfer (15%) is consistent with typical rates reported in inpatient settings.

20 participants had data available at both T1 and T2, and follow-up data (T3) were available for 14 participants. The six participants lost to follow-up (30%) reported relocation or lack of motivation as reasons for non-participation. The healthy control group comprised 12 participants assessed at T1 and T2; two children were excluded from T1 after developing mental symptoms. Groups were well-matched for age at baseline, with intervention (10.57 ± 1.75 years) and healthy control group (10.56 ± 1.41) nearly identical (Table 2). 10 individuals were enrolled in the wait-list control group (T0); no dropouts were recorded. The intervention group had a higher proportion of female participants (54%) compared to healthy controls (33%). Mean treatment duration was 7.91 (2.81) months, with follow-up assessments (T3) conducted 8.07 (1.66) months post-treatment.

**Table 2:** Baseline Characteristics and Descriptive Variables Across Study Groups and Timepoints. Data are presented as mean (standard deviation) for continuous variables or n (%) for categorical variables. Dashes (–) indicate missing data or variables that were not applicable. Abbreviations: **HCG**, Healthy Control Group; **IG**, Intervention Group; **WCG**, Waitlist Control Group; **T0**, Pre-treatment waitlist assessment; **T1,** Pre-treatment assessment; **T2**, Post-treatment assessment; **T3**, Follow-up assessment; **CBCL**, Child Behaviour Checklist; **SDQ**, Strengths and Difficulties Questionnaire; **WISC-IV,** Wechsler Intelligence Scale for Children, Fourth Edition; **PROPS**, Parent Report of Post-traumatic Symptoms; **SCARED,** The Screen for Child Anxiety Related Emotional Disorders; **DIKJ**, Depression Inventory for Children and Adolescents; **CDC**, Child Dissociative Checklist.

| Variable | Cut<br>off<br>(≥) | Total | HCG<br>(All) | IG<br>(All) | HCG<br>Pre<br>(T1) | HCG<br>Post<br>(T2) | WCG<br>Pre<br>(T0) | IG<br>Pre<br>(T1) | IG<br>Post<br>(T2) | IG Re<br>(T3) |
| --- | --- | --- | --- | --- | --- | --- | --- | --- | --- | --- |
| **N** | – | 85 | 24 | 61 | 12 | 12 | 10 | 27 | 20 | 14 |
| **Age** | – | 10.83<br>(1.71) | 10.80<br>(1.40) | 10.99<br>(1.78) | 10.56<br>(1.41) | 11.04<br>(1.41) | 9.90<br>(1.71) | 10.57<br>(1.75) | 11.15<br>(1.72) | 11.59<br>(1.83) |
| **Sex (female)** | – | 45<br>(47.37%) | 8<br>(33.33%) | 33<br>(54.10%) | 4<br>(33.33%) | 4<br>(33.33%) | 4<br>(40.00%) | 14<br>(51.85%) | 10<br>(50.00%) | 9<br>(64.29%) |
| **Handedness**<br>**(left)** | – | 14<br>(14.74%) | 2<br>(8.33%) | 10<br>(16.39%) | 1<br>(8.33%) | 1<br>(8.33%) | 2<br>(20.00%) | 4<br>(14.81%) | 3<br>(15.00%) | 3<br>(21.43%) |
| **CBCL** |  |  |  |  |  |  |  |  |  |  |
| Total | 38 | 44.84<br>(31.35) | 13.58<br>(8.80) | 52.21<br>(28.19) | 13.42<br>(7.08) | 13.75<br>(10.58) | 78.22<br>(25.40) | 67.59<br>(26.35) | 38.95<br>(24.99) | 41.50<br>(21.93) |
| Internalizing | 10 | 10.16<br>(8.21) | 3.29<br>(2.65) | 12.25<br>(8.24) | 3.33<br>(2.57) | 3.25<br>(2.83) | 14.33<br>(7.78) | 14.81<br>(8.36) | 10.40<br>(8.20) | 9.93<br>(7.10) |
| Externalizing | 17 | 18.03<br>(14.46) | 5.04<br>(4.34) | 20.93<br>(13.32) | 4.75<br>(3.82) | 5.33<br>(4.96) | 33.00<br>(14.98) | 27.85<br>(13.06) | 14.25<br>(11.31) | 17.14<br>(10.33) |
| **CBCL T scores** |  |  |  |  |  |  |  |  |  |  |
| Total | 64 | 63.65<br>(11.76) | 50.08<br>(6.08) | 67.25<br>(9.54) | 50.17<br>(5.27) | 50.00<br>(7.03) | 75.44<br>(3.50) | 72.15<br>(7.93) | 62.60<br>(9.84) | 64.43<br>(7.81) |
| Internalizing | 64 | 60.06<br>(10.42) | 50.33<br>(6.46) | 62.98<br>(9.54) | 50.67<br>(6.23) | 50.00<br>(6.94) | 66.22<br>(8.20) | 66.07<br>(8.77) | 60.55<br>(10.04) | 60.50<br>(9.15) |
| Externalizing | 64 | 62.87<br>(12.24) | 49.71<br>(7.31) | 66.44<br>(10.20) | 49.42<br>(7.15) | 50.00<br>(7.77) | 73.78<br>(7.55) | 71.48<br>(8.36) | 60.95<br>(10.72) | 64.57<br>(8.33) |
| **SDQ** |  |  |  |  |  |  |  |  |  |  |
| Total | 17 | 14.11<br>(7.78) | 5.50<br>(4.17) | 16.43<br>(6.30) | 5.50<br>(3.03) | 5.50<br>(5.21) | 21.33<br>(5.87) | 18.85<br>(5.16) | 14.30<br>(6.42) | 14.79<br>(6.91) |
| Hyperactivity | 7 | 5.10<br>(2.93) | 2.54<br>(2.43) | 5.77<br>(2.62) | 2.83<br>(2.37) | 2.25<br>(2.56) | 7.33<br>(1.58) | 6.48<br>(2.50) | 5.15<br>(2.68) | 5.29<br>(2.61) |
| **WISC-IV** |  |  |  |  |  |  |  |  |  |  |
| Total | – | – | – | – | – | – | – | 92.73<br>(18.46) | – | – |
| Working<br>Memory | – | 98.55<br>(16.55) | 108.50<br>(15.08) | 95.36<br>(16.47) | 107.75<br>(16.75) | 109.25<br>(13.92) | 91.89<br>(8.70) | 92.82<br>(15.29) | 96.15<br>(18.64) | 98.21<br>(15.53) |
| **Trauma**<br>**(PROPS)** | 17 | 16.21<br>(12.17) | 3.79<br>(4.66) | 19.34<br>(10.65) | 3.17<br>(3.21) | 4.42<br>(5.85) | 28.11<br>(10.55) | 24.78<br>(9.17) | 14.30<br>(10.96) | 16.07<br>(8.21) |
| **Anxiety**<br>**(SCARED)** | 25 | 12.59<br>(9.79) | 6.04<br>(6.04) | 14.51<br>(9.74) | 6.50<br>(7.00) | 5.58<br>(5.18) | 17.00<br>(10.89) | 15.67<br>(10.96) | 13.65<br>(8.92) | 13.50<br>(8.71) |
| **Depression**<br>**(DIKJ)** | 15 | 11.27<br>(8.66) | 6.64<br>(2.87) | 12.18<br>(9.23) | 6.60<br>(3.78) | 6.67<br>(2.02) | 17.62<br>(9.83) | 12.55<br>(10.50) | 12.65<br>(9.97) | 10.93<br>(5.92) |
| **Dissociation**<br>**(CDC)** | 12 | 9.66<br>(5.69) | – | 9.41<br>(5.68) | – | – | 12.33<br>(4.90) | 11.19<br>(5.71) | 8.15<br>(6.17) | 7.79<br>(4.00) |

### Clinical outcomes

At baseline, the intervention group demonstrated clinically elevated symptoms across most measures (Table 2), with mean scores exceeding established clinical cutoffs for nearly all variables. In contrast, healthy controls consistently scored in the non-clinical range across all timepoints. Following treatment, the intervention group showed significant symptom reduction across multiple domains. Mixed-effects analyses revealed significant group-by-time interactions for CBCL total problems (F(1, 58.53) = 4.72, p = .034, ω²□ = .058), externalizing behaviour (F(1, 57.37) = 7.00, p = .011, ω²□ = 0.092) and trauma symptoms (F(1, 59.86) = 3.93, p = .048, ω²□ = .051). Post-hoc analyses demonstrated large effect sizes for changes from baseline to post-treatment across multiple domains (Table S3). Several measures achieved remission below clinical cutoffs, including externalizing behaviour (d = -1.79, CI95% [-1.07, -2.51], p < .001), SDQ total difficulties (d = -1.37, CI95% [-0.68, -2.07], p < 0.001) and trauma symptoms (d = -1.19, CI95% [-0.51, -1.86, p = .005) and hyperactivity (d = -1.23, CI95% [-0.54, -1.92], p = .004). CBCL total problems showed significant improvement with a large effect size (d = -1.84, CI95% [-1.12, -2.56], p < .001) but remained in the subclinical range. Treatment gains were maintained at six-month follow-up, with overall effect sizes remaining large (dmean = -1.10 compared to baseline). A slight non-significant increase in some symptom scores was observed between post-treatment and follow-up (d_mean_ = 0.01), particularly for CBCL total problems (d = 0.19, CI95% [0.19 -0.93], p = .602), externalizing behaviour (d = 0.40 , CI95% [1.15, -0.34], p = .784) and trauma symptoms (d = 0.18, CI95% [0.89, -0.54], p = .956), though scores remained significantly below baseline levels. This pattern is commonly observed and likely at least in part due to the transition from a highly structured inpatient treatment to often suboptimal home and school environments.

Internalizing symptoms (d = -0.77), depression (d = -0.20), and anxiety (d = -0.44) showed medium to small treatment responses and effect sizes.

The healthy control group demonstrated no significant symptom changes, highlighting the specificity of treatment effects. The course of symptoms for the intervention and the control group is visualised in Figure 2.

**Figure 2:**
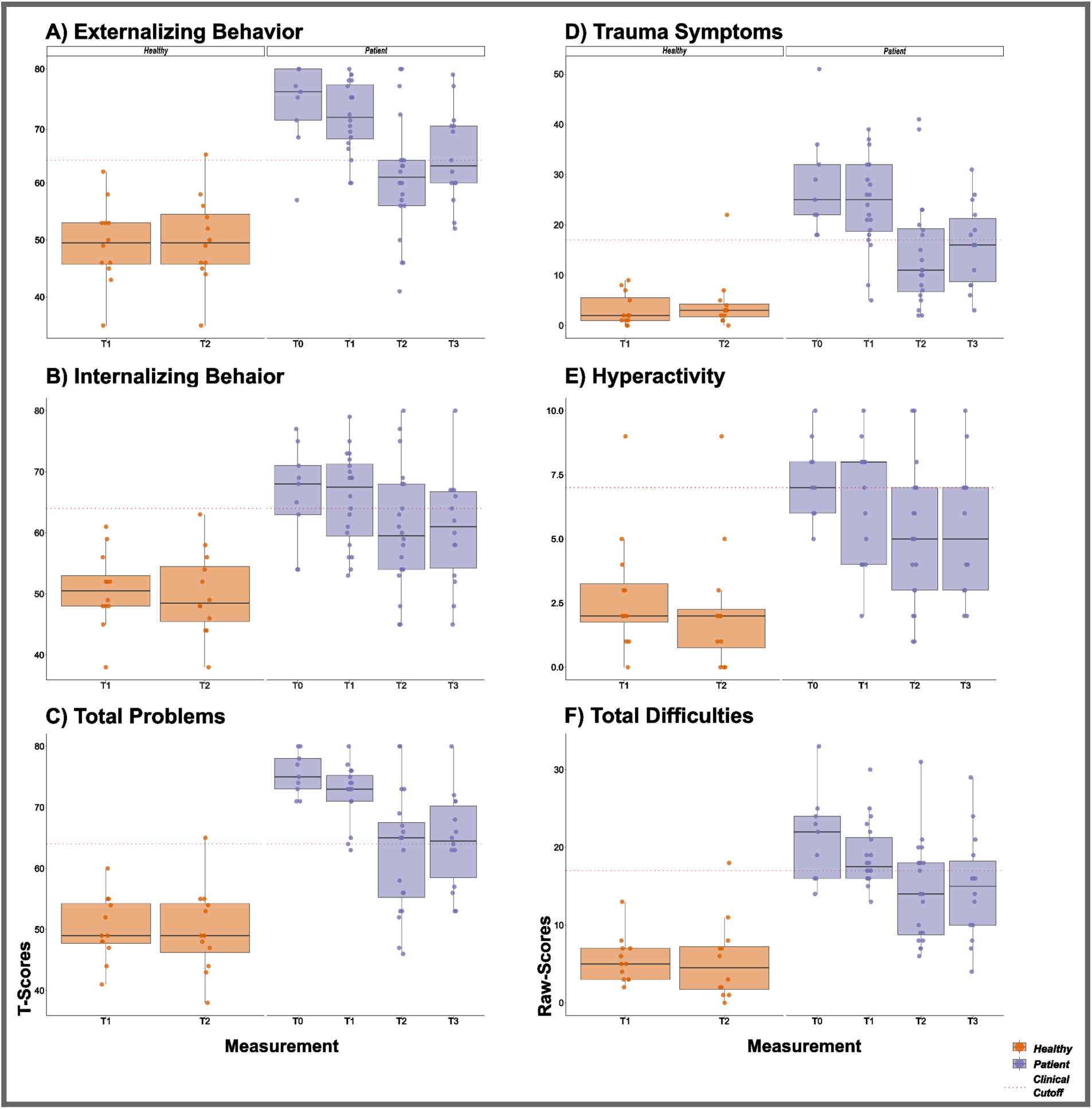
Overview on Main Clinical Progress. Scatter-boxplots illustrate the distribution of symptom scores across the study timeline for the Patient Cohort (Purple) and Healthy Controls (Orange). Clinical factors are presented, including t-scores from the Child Behaviour Checklist (CBCL) across the following domains represented as scatter-boxplots: A) Externalizing Behaviour, B) Internalizing Behaviour, and C) Total Problems. Trauma symptoms are assessed using D) Parent Report of Posttraumatic Stress Symptoms (PROPS), along with E) Hyperactivity and F) Total Difficulties from the Strengths and Difficulties Questionnaire (SDQ). Scores are displayed for patients in the Waitlist Control Group (T0) and the Intervention Group (T1 = baseline, T2 = post-treatment, T3 = follow-up) as well as healthy controls (T1, T2), with the clinical cutoff indicated by a red dotted line.

**Figure 3:**
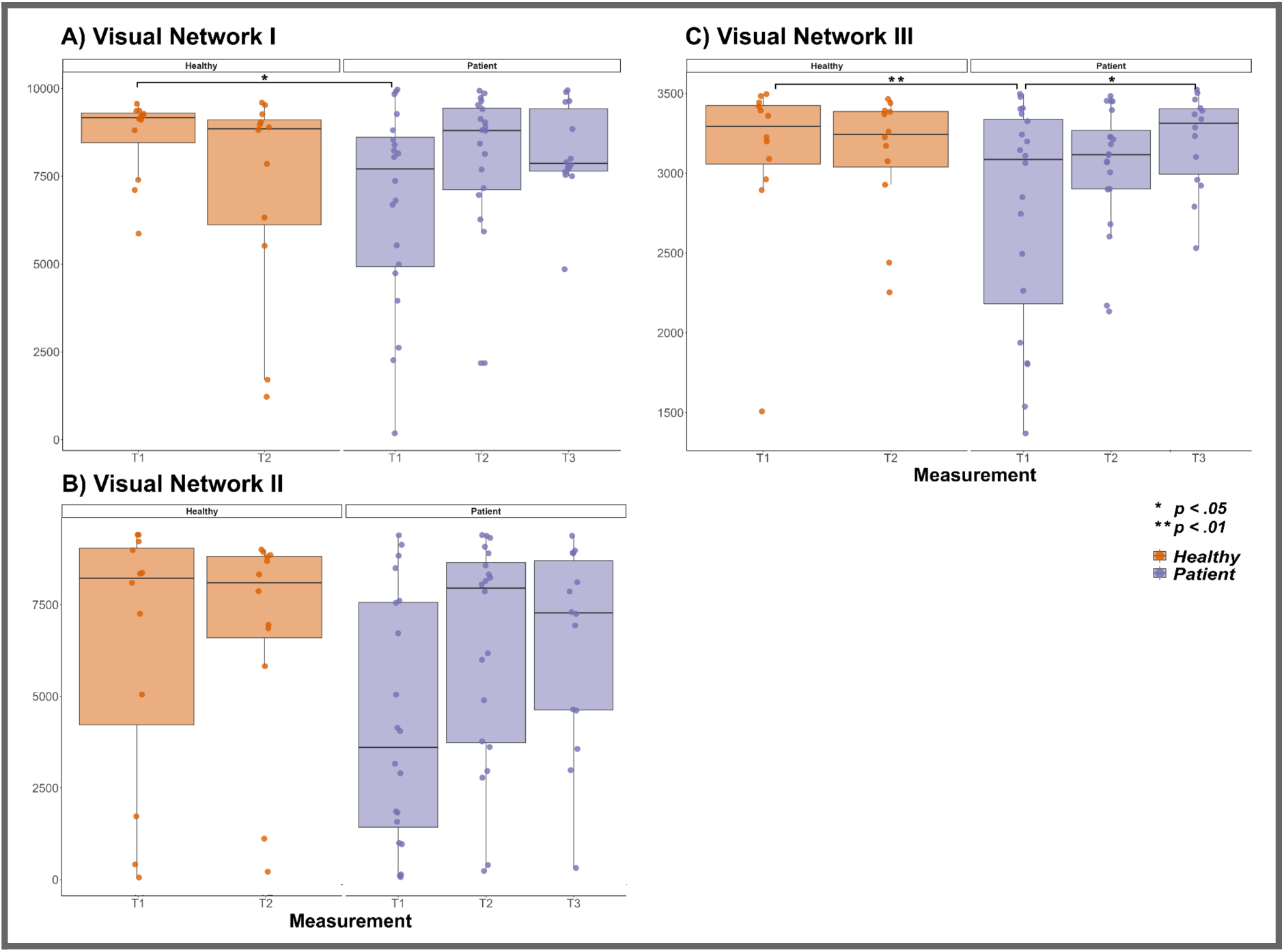
Longitudinal Functional Reorganization of Visual Processing Networks. Spatial extent of activated voxels within three distinct visual networks (threshold: z > 4) are displayed for A) Visual Network I, B) Visual Network II, and C) Visual Network III. The plots compare the trajectories of the Intervention Group (Purple) across Baseline (T1), Post-Treatment (T2), and Follow-Up (T3) against the stable baseline of Healthy Controls (Orange) at T1 and T2. Note: The intervention group generally exhibits a pattern of lower activation at baseline increasing toward healthy control levels post-treatment.

Regarding dropouts, there was no relationship to baseline symptom load. For example, in terms of CBCL total and depression scores, the 7 dropouts at T1 had the same distribution as the complete cohort. The 6 dropouts at T2 were all below the clinical threshold in terms of CBCL total scores, suggesting that it’s unlikely they did not return for follow-up due to dissatisfaction with post-treatment results and aligned with the reasons they reported (relocation and lack of motivation).

### Neuroimaging outcomes

#### Specific Reorganization of Visual Processing Networks

The most striking effects emerged in visual processing networks, where the intervention group showed distinct patterns of change following treatment. In Visual Network I, significant group-by-time interactions were observed (F(1, 51,16) = 9.20, p = .004, ω²□ = .132). At baseline, the intervention group showed significantly lower activation compared to healthy controls (d = -1.38, CI95% [-0.45, -2.31], p = .030), but the difference disappeared following treatment (p = .796). Within the intervention group, activation increased from baseline to post-treatment (d = 0.55, CI95% [1.21, -0.11], p = .442) and continued to increase through follow-up (d = 0.67, CI95% [1.45, -0,11], p = .428), representing a pattern of normalization toward healthy control levels. Similar patterns emerged in Visual Networks II and III.

Visual Network II showed marginally significant group differences (F(1, 52,34) = 3.45, p = .069, ω²□ = .069) and group-by-time interactions (F(1, 50.67) = 3.28, p = .076, ω²□ = .041).

Visual Network III provides further evidence for substantial group differences (F(1, 52,34) = 4.97, p = .030, ω²□ = .059) and approaches significance for parameters of both time (F(2, 52.04) = 2.80, p = .070, ω²□ = .053) and the interaction between group-by-time (F(1, 51,19) = 3.68, p = .061, ω²□ = .04). The intervention group shows significant activation increases from baseline to follow-up (d = 1.17, CI95% [1.96, 0.37], p = .033). While group contrasts were evident at baseline (d = -1.59, CI95% [-0.65, -2.52], p = .007), they vanished by the time of post-testing (d = -0.40, CI95% [-0.40, 0.47], p = .717). Within-subject testing further underscores temporal changes from baseline to follow-up (d = 1.17, CI95% [1.96, 0.37], p = .033 ).

#### Specificity of Effects and Confounding Factors

Results concerning the Default Mode Network - subdivided into frontomedial (F(1, 52.34) = 3.10, p = .084, ω²□ = .032) and posterior components (F(1, 52.35) = 8.59, p = .005, ω²□ = .032) - revealed significant baseline differences across both with small effect sizes. However, substantial motion-related noise was detected in both ICs, and post hoc analyses did not support a treatment-related association.

Conservative analysis of potential confounding factors (p < 0.1) revealed strongest effects of headmotion on following components: Somatomotor Network (F(1, 59.52) = 9.56, p = .003, ω²□ = .128), Default Mode Network (F(1, 63.48) = 9.06, p = .004, ω²□ = .116 ) and the fronto-medial division of the Default Mode Network Network (F(1, 59.36) = 7.10, p = .010, ω²□ = 0.090). Minor effects of head motion were observed in the left-lateralized fronto-parietal control network approaching statistical significance (F(1, 62.61) = 3.74, p = .058, ω²□ = .044).

Critically, the visual networks that emerged as most treatment-responsive were largely unaffected by motion artifacts. Nonetheless, significant effects were observed for Visual Network I associated with confounders handedness (F(1, 21.75) = 6.06, p = .022, ω²□ = .163) and gender (F(1, 31.51) = 4.85, p = .035, ω²□ = .097), and for Visual Network III with the interaction between group and scanner type (F(1, 50.23) = 4.14, p = .047, ω²□ = .048), as well as with the presence of a companion (parent or medical staff) during scan acquisition (F(1, 62.80) = 2.81, p = .068, ω²□ = .056).

Differences for visual and other networks are summarised in Table S4.

### Associations between neuroimaging and clinical outcomes

Healthy controls demonstrated large negative correlations between visual network activation and total symptom severity (r(10) = -.79, CI95% [-.94, -.39], p = .002), hyperactivity (r(10) = -.86, CI95% [-.96, -.57], p < .001) and externalizing behaviour (r(10) = -.67, CI95% [-.90, -.15], p = .018), presumably consistent with efficient processing (see Table S5 for full correlation parameters). In contrast, the intervention group showed moderate to large positive correlations at baseline for total symptom severity (r(24) = .36, CI95% [-.03, .65], p = .073), hyperactivity (r(24) = .43, CI95% [.05, .70], p = .030) and externalizing behaviour (r(24) = .40, CI95% [.02, .68], p = .041), suggesting that higher visual network activity was associated with greater symptom load(detailed correlation metrics for this cohort are provided in Table S6). Following treatment, these possibly maladaptive positive correlations disappeared in the intervention group for total symptom severity (r(18) = -.01, CI95% [-.45, .44], p = .975), externalizing behaviour (r(18) = -.12, CI95% [-.53, .34], p = .626) and hyperactivity (r(18) = .24, CI95% [-.22, .66], p = .305), with brain-behaviour relationships approximating those observed in healthy controls and remained stable at follow-up. This pattern of correlation changes was consistent across multiple visual networks and clinical measures, but was absent when looking at the correlations of other networks with clinical measures.

This suggests that efficient visual network integration might support adaptive behavioural regulation. For example, activity in Visual Network III negatively correlated with both hyperactivity (r(10) = -.86, CI95% [-.96, -.57], p < .001) and total difficulties (r(10) = -.79, CI95% [-.94, -.39], p = .002) in healthy peers. Conversely, at baseline (T1), highly traumatized children exhibited a paradoxical, maladaptive pattern. Higher activation in Visual Networks I and II significantly correlated with greater symptom severity, including increased dissociation (r (24) = .54, CI95% [.19, .77], p = .004) and hyperactivity (r (24) = .46, CI95% [.09, .72], p = .019). Following intensive psychotherapy, this maladaptive coupling was effectively reorganised. At post-treatment (T2), the pathological positive correlations between visual network activity and clinical symptoms disappeared. By follow-up (T3), the relationship began to approximate the healthy profile. For instance, higher activity in Visual Network III predicted lower anxiety (r (14) = -.57, CI95% [-.83, -.11], p = .020).

## 5. Discussion

This study provides the first evidence that intensive, medication-free inpatient treatment can achieve clinical remission and neurobiological normalisation in severely traumatized children. Our primary hypothesis, that intensive trauma-informed treatment would reduce behavioural and affective self-regulation difficulties was supported, with participants achieving significant improvements in externalizing behaviours and trauma symptoms, that were attenuated but sustained at 6-month follow-up. Furthermore, brain-behaviour coupling patterns for visual networks normalised to resemble those observed in healthy controls. These findings demonstrate that even in children with severe complex trauma histories and multiple placement failures, intensive psychotherapeutic intervention without pharmacological support can achieve both clinical remission and changes in neural function.

The literature around trauma-informed interventions for children and adolescents is growing. Given the diversity of published treatment approaches, settings and cohorts, a precise comparison is necessary and helps clarify what is unique to our study and treatment approach:

1. While most trauma-informed studies take place in “residential settings” or in the context of “youth services”, dedicated psychiatric inpatient settings are reported considerably rarer. Children treated in our cohort had already experienced multiple placement and service failures due to an inability to function in group settings, thus the intensive inpatient treatment was a last resort.
2. Most published studies either do not comment on the use of medication or used medication as an adjunct treatment. For example, one study showed that 40% of participants were on medication at the beginning of their study and 18% at the end^57^, while in our case, 100% were on (often multiple) medications that were discontinued before the study began.
3. To meet the deep clinical need of our children without the use of medication, our program was much more intensive than what is typically reported under “trauma-informed interventions”. Published approaches at the lower end of intensity can consist of “educational meetings”^25^ or group formats employing uniform, evidence-based treatment protocols consisting of a few sessions mostly tailored for outpatient interventions like TF-CBT^12–15^ or EMDR^16–18^, while our program consisted of almost daily psychotherapy sessions, daily staff reflection sessions and a time-intensive attachment regulation approach (with 1:1 or 2:1 support) inside the milieu therapeutic group framework.
4. Finally, to the best of our knowledge, this is the first study presenting longitudinal neuroimaging in the context of such an intensive treatment program for severely traumatized children.

Comparisons with other studies are inherently fraught due to the often unique circumstances in RTCs^23^, the different inclusion criteria and symptom profiles manifesting in different age ranges, the wide variety of assessment tools and treatments utilised, the overall sparseness of studies and the quality concerns in many of those published^58^. Nonetheless, some tentative comparisons can be made. First, the use of psychodynamic therapy in this patient population is an exception; most published studies report the use of CBT and supplementary therapies, like outdoor-based interventions^58^. Furthermore, we have not found any study that offered comparable treatment intensity and duration. Many RCTs offer short-term treatments and lean heavily on group therapies^58^, while the approach implemented in this study was centred around almost daily individual psychotherapy sessions.

Beyond intensity differences, the approach outlined in this article fundamentally reconceptualizes core treatment modules. For example, where standard residential treatment relies on case-handling by rotating staff, we provide designated consistent primary therapist/caregiver/physician as attachment figures throughout six-to-eight-months stays on average, with multiple weekly support sessions for the attachment figures.

As for treatment effects, while most studies show symptom improvement post intervention in some domains, the magnitude of improvement and the baseline severity in our cohort stands out. For example, in one study, 43 female youths in residential care with history of sexual abuse received group therapy and 60% of participants received individual therapy^59^. Six months of CBT and trauma-informed therapy (comparable to our treatment duration) reduced the overall CBCL score from 67.3 to 64.4. Another well-powered study (n = 126) employed a trauma-informed treatment and saw a very similar reduction in CBCL total score from 67.5 to 64^60^. In comparison, our baseline and the reduction in our cohort was much more substantial from 72.2 to 62.6.

A recent RCT study had a very similar age range to our cohort (7-13 years) and looked at externalising symptoms after the augmentation of pre-existing relationally-based treatment with a novel approach developed for youth with trauma history^61^. While the augmentation produces a notable improvement (Cohen’s d 0.35 before and 0.7 after implementation), this remained significantly below the effect sizes observed in our cohort (d = 1.78). The observed large effect size for externalising symptoms is of particular relevance, as most published studies focus on internalizing related outcomes^58^, which is comprehensible, given that highly burdened subjects are difficult to research if they lack basic group competence, exhibit low impulse control and show excessive disruptive behaviour. Yet a recent meta-analysis revealed that the pooled prevalence of externalising problems is significantly higher, especially in children in residential care (49% vs. 39%)^62^. The more dysregulated the behaviour of the children presents, the heavier treatment tends to lean on time-out placements (TOPs), which are employed in roughly 33% of residential settings serving children and adolescents exhibiting severe externalizing behaviour. TOPs are intended as short-term environmental removals to interrupt behavioural escalation with the goal of consecutive reintegration into the original environment^29^. However, in reality TOPs are often implemented under conditions of staff exhaustion and institutional strain, conditions which usually accompany such a therapeutically challenging sample. Interviewed staff often admits systemic issues with TOPs, describing prison-like transport and vague institutional objectives^63^.

Therefore, they are thought to at times re-traumatize individuals, leading to further TOP interventions as well as restraint and service disengagement^29^. The implementation of trauma-informed care has been shown to significantly reduce the incidents of physical restraint^60^, with some studies reporting reductions in seclusion room incidents^64^. However, the way how the restraint is performed might be of equal importance to its frequency. Our approach makes use of a “time-intensive” method, where restraint is coupled with intensified attachment regulation practices in situations of behavioural escalation. Instead of being isolated, the children were accompanied out of the situation on a 2:1 caregiver:patient basis and received emergency psychotherapy if required.

Another very common way behavioural dysregulation is managed is the use of pharmacology. However, most medications (aside from a few stimulants and antipsychotics) demonstrate only small to medium effect sizes^65^. Furthermore, the clinical use of medications should be weighed against substantial side-effect profiles, ranging from acute insomnia and appetite suppression to more systemic risks such as metabolic changes and growth inhibition^26,27^. Beyond physiological side effects, pharmacological intervention may increase acute safety risks, such as higher rates of non-suicidal self-injury^66^. Of particular concern are pharmacological impacts on the developing brain. These medications are intended to target dysregulated neural circuits; the goal is to achieve functional and structural normalisation^67^. The developing brain’s concurrent structural remodelling and neurochemical maturation increase the susceptibility to drug exposure, particularly regarding the impact of psychostimulants on central nervous system development^28^. The heightened sensitivity necessitates pediatric pharmacological research to determine how such treatments might fundamentally alter normative growth trajectories and long-term neurobiological outcomes^68,69^.

Given these issues, our large and sustained symptom improvements without pharmacological intervention highlight the potential of intensive trauma-informed psychotherapy, even in severely dysregulated children.

Beyond clinical outcomes, this study provides novel evidence that intensive trauma-informed care is associated with functional reorganization of neuronal networks in highly traumatized children. The most pronounced and consistent effects were observed in visual processing networks, which showed baseline alterations relative to healthy controls and progressive normalisation following treatment.

These findings align with developmental models of maltreatment-related brain adaptation, where maltreatment alters trajectories of brain development to affect sensory systems, network architecture, and circuits involved in threat detection. Specifically, alterations in visual processing areas (such as V1 and V2) have been identified as adaptive modifications to facilitate the rapid detection of threat in malevolent environments^8,33^. In line with these findings, prior structural neuroimaging research has consistently documented maltreatment-related gray matter volume reductions and cortical thinning in the primary and secondary visual cortices^31,32^. Additionally, meta-analytic evidence highlights widespread microstructural abnormalities in the visual-limbic white matter tracts, such as the inferior longitudinal fasciculus and optic radiations, which process and convey adverse visual input^31,70^. However, it is unknown whether these sensory adaptations can be reorganized through therapeutic experiences. Our results support this hopeful hypothesis, showing that the intervention promoted a normalisation of networks involved in sensory processing and threat detection, bringing them into closer alignment with those observed in healthy controls. While these represent exploratory results, the convergence of clinical remission with neuroimaging changes is compelling, suggesting that treatment effects might involve fundamental reorganization of brain-behaviour relationships rather than simple symptom reduction.

The present results suggest that intensive therapeutic environments - characterized by safety, predictability, and relational attunement - may support recalibration of early sensory systems. Importantly, effects were network-specific and not globally distributed across all canonical networks, arguing against nonspecific maturation or scanner-related explanations.

A particularly compelling finding is the observed shift in brain-behaviour relationships. At baseline, traumatized children exhibited paradoxical positive associations between visual network activity and symptom severity, suggesting maladaptive coupling between sensory processing and behavioural regulation. Following treatment, these maladaptive correlations dissipated and approximated the inverse relationships observed in healthy controls.

This “flipping” of brain-behaviour coupling - from maladaptive to normalized patterns- suggests that clinical improvement may reflect restoration of functional integration rather than mere symptom reduction. Such coupling metrics may represent promising translational markers of treatment response, complementing traditional symptom-based outcomes.

Our study is unique in its inclusion of highly traumatized children who experienced multiple placement failures and its exclusive reliance on intensive, non-pharmacological inpatient treatment. Nonetheless, several limitations should be acknowledged. While the intensive inpatient treatment showed very promising results, it was not evaluated in a randomized controlled trial, so causal conclusions remain tentative. The very resource-intensive nature of this intervention presents a challenge for large-scale research, which is evident in our limited sample size and limited long-term follow-ups. Despite these constraints, the findings generate valuable hypotheses that future multi-site, adequately powered RCTs with longer follow-up periods should examine.

Taken together, these findings suggest that children exposed to cumulative interpersonal trauma may benefit from treatment approaches that go beyond standard outpatient PTSD protocols. Intensive, trauma-informed inpatient psychotherapy may engage fundamental neurodevelopmental mechanisms, particularly within early sensory systems, supporting both clinical recovery and normalisation of brain-behaviour relationships.

## Data Availability

All data produced in the present study are available upon reasonable request to the authors

## 6. Acknowledgements

The authors thank the patients, parents and other primary caregivers, as well as the children and their parents in the healthy control group for their participation. We are very grateful to the nursing and pedagogical staff, teachers, psychotherapists, physicians, art, music, movement and sports therapists for their strong commitment to treating these severely traumatized children and adolescents. We thank our external supervisors, without their support we could not have provided this intensive inpatient treatment. We also thank the research assistants for data collection, ratings, evaluation and data preparation for statistical analyses.

## Author contribution

K.H.B. and J.Q. designed and conceived the clinical study protocol, with K.H.B. additionally designing the intervention model. K.H.B. acquired the grant and secured funding for the study. D.K. and B.E.W. designed the neuroimaging study and supervised all aspects. B.E.W. evaluated all clinical scans, D.K. was involved in all pre- and post-neuroimaging steps and supervised L.E. in the specific evaluation steps and quality control. C.D., L.O., A.B., L.E., J.G. and K.H.B. were involved in patient recruitment and clinical data collection, with C.D., L.O., L.E., J.G. and A.B. performing all aspects of the clinical tests. J.G. performed individual therapies, and L.E. performed group and individual therapies. The neuroimaging procedures and evaluations were supervised by D.K. and B.E.W. K.H.B. was responsible for data monitoring and F.P. for clinical monitoring with regard to traumatization. The statistical analyses were performed by L.E. under the statistical supervision of S.G., M.K. and D.K. M.K. wrote the draft manuscript with the participation of L.E., L.Y., M.M., V.W. and D.K. M.K., L.E., M.M., L.Y., V.W., M.K., S.G., S.S., F.P., B.E.W., K.H.B., and D.K. discussed the results, provided critical comments, and made corrections to the manuscript. All authors reviewed the manuscript prior to submission.

## Supporting Material

**Table S1:** Clinical Assessment and Study Enrollment Protocol. Overview of the sequential appointments comprising the initial clinical contact, diagnostic interviewing, and multi-modal assessments (cognitive, psychological, and neurobiological) conducted prior to treatment. Abbreviations: **DIKJ**, Depression Inventory for Children and Adolescents.

| **Appointment** | **Purpose** | **Notes** |
| --- | --- | --- |
| Appointment 1 | Parent consultation (child not present) | Team composition: Research coordinator, inpatient unit representative, psychotherapist, attending physician. |
| Appointment 2 | Child introduction & clinical interview | Participants: Child, psychotherapist, attending physician.<br>Components: Meet and greet, clinical interview with parents and child to discuss presenting concerns. |
| Appointment 3 | Cognitive & medical assessment | Intelligence testing administered by psychotherapist and physical examination. |
| Appointment 4 | Projective assessment & family interaction | Projective measures: Familie in Tieren, Baum Test, Scenotest.<br>Dyadic play observation (child with both parents). |
| Appointment 5 | Attachment assessment | DIKJ (administered first), attachment interview. |
| Appointment 6 | Neuroimaging | NA |
| Appointment 7 | Feedback & treatment planning session | NA |

**Table S2:** Functional Magnetic Resonance Imaging (fMRI) Baseline Acquisition and Motion Parameters. Distribution of MRI scanner types (Verio vs. Skyra) and quantitative in-scanner head motion parameters (absolute and relative mean displacement) for the Intervention and Healthy Control groups across baseline (T1), post-treatment (T2), and follow-up (T3) sessions. *Note: *mean (SD) or n (%)*.

|  | Total | Healthy Control Group (HCG<sub>ALL</sub>) | Intervention Group (IG<sub>All</sub>) | Healthy Control Group (HCG) |  | Intervention Group (IG) |  |  |
| --- | --- | --- | --- | --- | --- | --- | --- | --- |
|  |  |  |  | Pre (T1) | Post (T2) | Pre (T1) | Post (T2) | Re (T3) |
| *N* | 85 | 24 | 61 | 12 | 12 | 27 | 20 | 14 |
| **Scanner type** |  |  |  |  |  |  |  |  |
| Skyra | 26 (30.59%) | 14 (58.33%) | 12 (19.67%) | 4 (33.33%) | 10 (83.33%) | 2 (7.41%) | 5 (25.00%) | 5 (35.71%) |
| Verio | 59 (69.41%) | 10 (41.67%) | 49 (80.33%) | 8 (66.67%) | 2 (16.67%) | 25 (92.59%) | 15 (75.00%) | 9 (64.29%) |
| **FMRI-motion** |  |  |  |  |  |  |  |  |
| absolute | 0.61 (0.62) | 0.53 (0.58) | 0.64 (0.64) | 0.65 (0.80) | 0.42 (0.22) | 0.70 (0.82) | 0.70 (0.82) | 0.42 (0.27) |
| relative | 0.13 (0.16) | 0.13 (0.17) | 0.14 (0.16) | 0.16 (0.22) | 0.09 (0.09) | 0.14 (0.19) | 0.14 (0.19) | 0.12 (0.12) |
| Handedness (left) | 12 (14.12%) | 2 (8.33%) | 10 (16.39%) | 1 (8.33%) | 1 (8.33%) | 4 (14.81%) | 3 (15.00%) | 3 21.43%) |

**Table S3:**
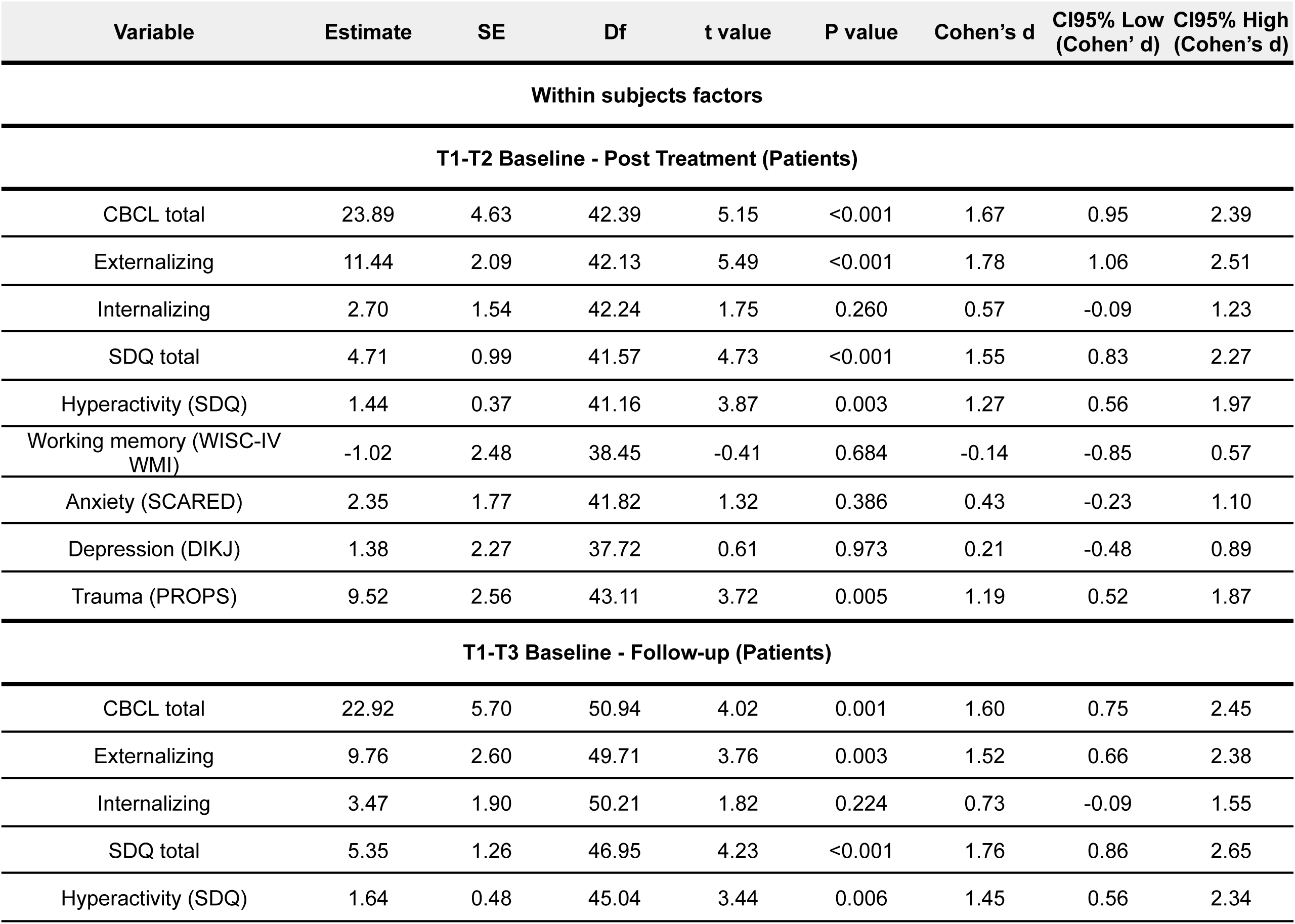
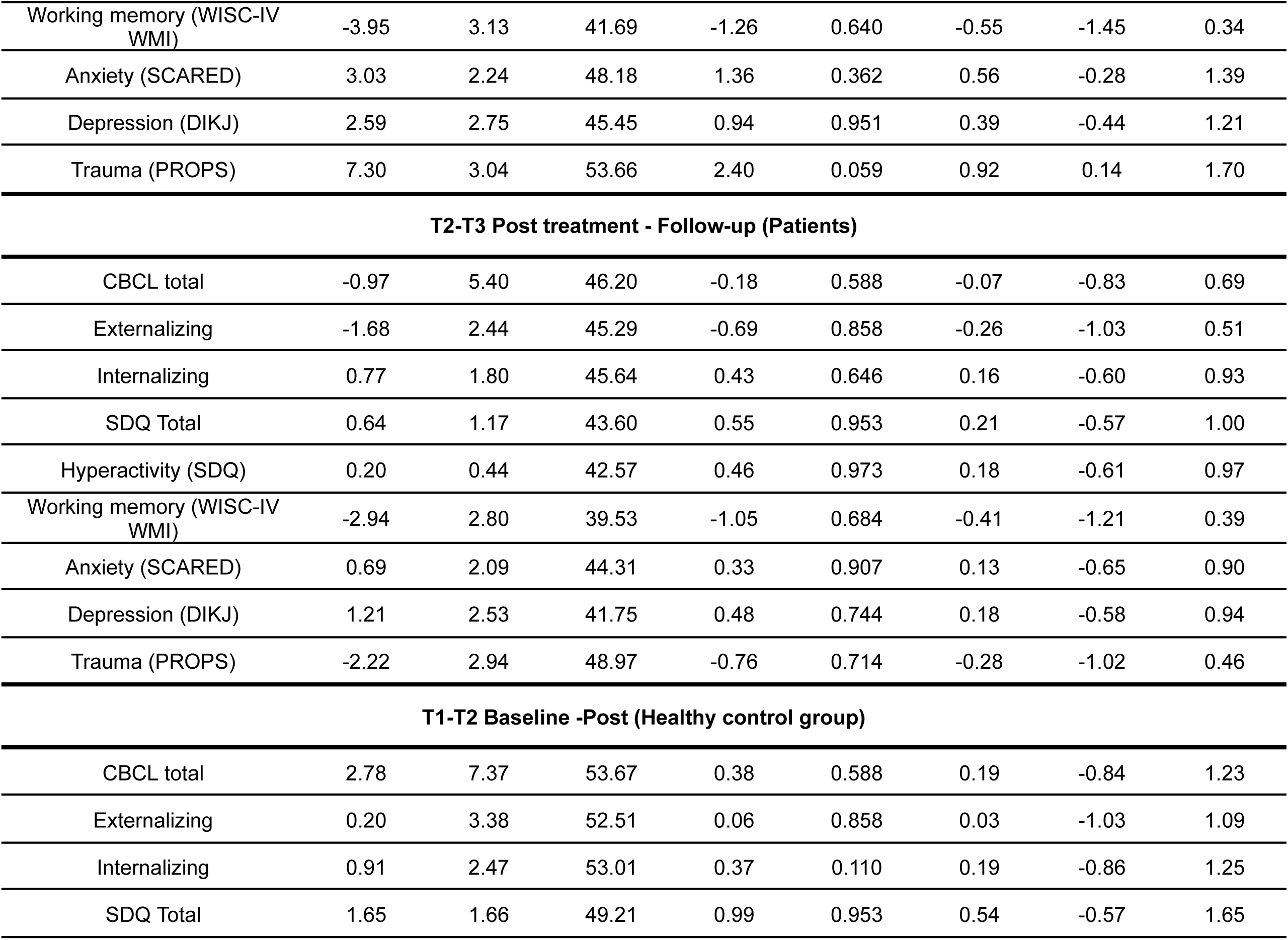
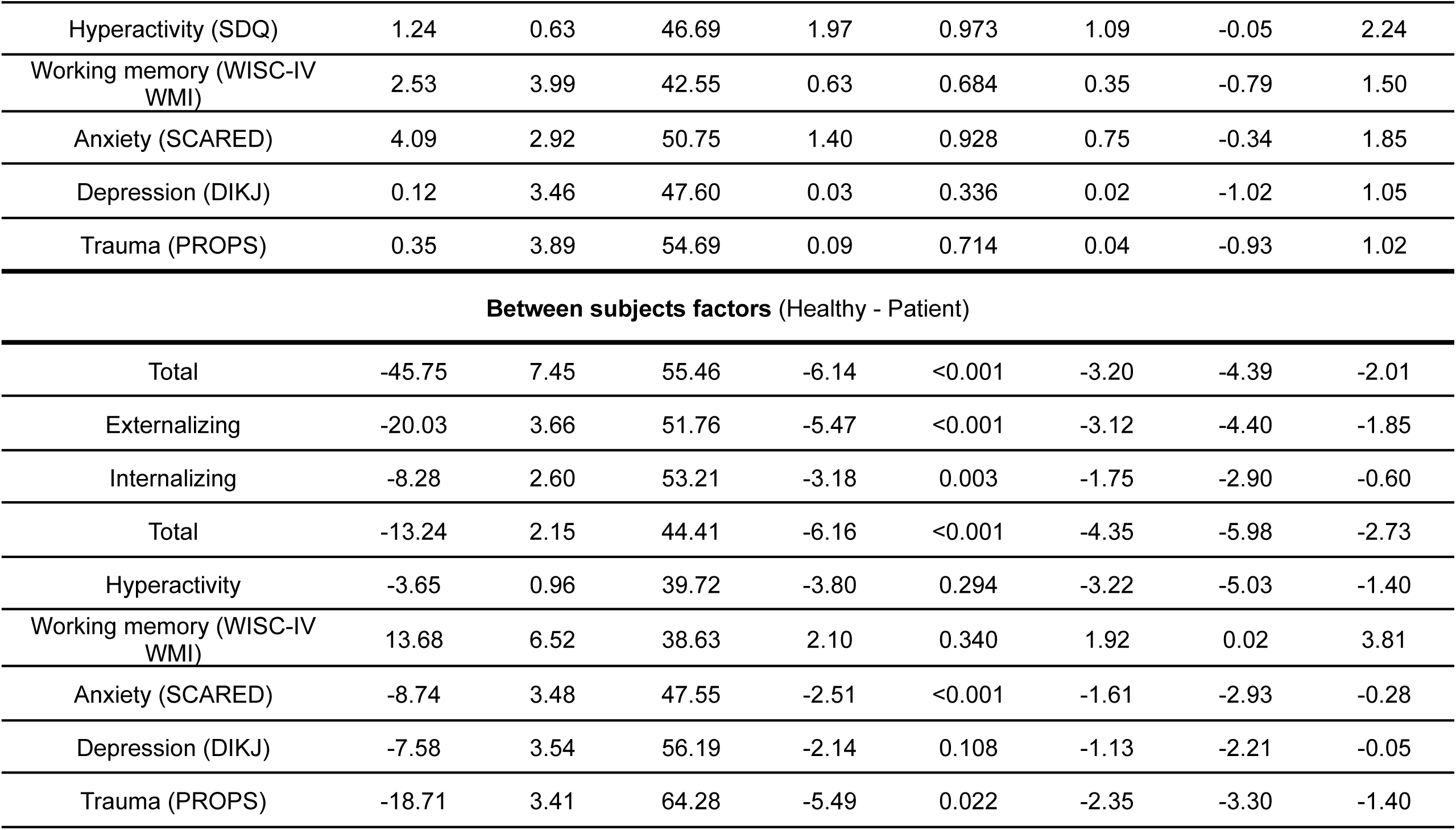
Post-hoc Pairwise Comparisons of Clinical Variables. SE standard error, Df Kenward-Roger adjusted degrees of freedom, P values were Hommel-adjusted, Cohen’s d was reported for both within-subject (longitudinal) and between-subject (group) contrasts. Abbreviations: **CBCL**, Child Behaviour Checklist; **SDQ**, Strengths and Difficulties Questionnaire; **WISC-IV,** Wechsler Intelligence Scale for Children, Fourth Edition; **PROPS**, Parent Report of Post-traumatic Symptoms; **SCARED,** The Screen for Child Anxiety Related Emotional Disorders; **DIKJ**, Depression Inventory for Children and Adolescents; **CDC**, Child Dissociative Checklist.

**Table S4:** Post-hoc Pairwise Comparisons of Functional Network Activation. Detailed statistical results of post-hoc pairwise. t-tests evaluating the spatial extent of activated voxels within specific resting-state networks (threshold z>4). SE standard error, Df Kenward-Roger adjusted degrees of freedom, P values were Hommel-adjusted, Cohen’s d was reported for both within-subject (longitudinal) and between-subject (group) contrasts.

| Variable | Estimate | SE | Df | t value | P value | Cohen's d | CI95% Low<br>(Cohen's d) | CI95% High<br>(Cohen's d) |
| --- | --- | --- | --- | --- | --- | --- | --- | --- |
| Within subjects factors |  |  |  |  |  |  |  |  |
| T1-T2 Baseline - Post Treatment (Patients) |  |  |  |  |  |  |  |  |
| Visual I | -1204.48 | 710.24 | 41.70 | -1.70 | 0.442 | -0.55 | -1.21 | 0.11 |
| Default mode (frontomedial) | 75.20 | 930.09 | 41.79 | 0.08 | 0.936 | 0.03 | -0.62 | 0.68 |
| Motor | 27.04 | 1043.12 | 41.79 | 0.03 | 0.979 | 0.01 | -0.64 | 0.66 |
| Visual II | -2159.89 | 945.48 | 41.12 | -2.28 | 0.221 | -0.75 | -1.42 | -0.08 |
| Salience | 149.15 | 359.76 | 41.51 | 0.41 | 0.973 | 0.14 | -0.52 | 0.79 |
| Visual III | -337.97 | 156.19 | 41.79 | -2.16 | 0.217 | -0.70 | -1.37 | -0.04 |
| Dorsolateral frontoparietal<br>(attention) | -310.61 | 275.13 | 41.14 | -1.13 | 0.894 | -0.37 | -1.03 | 0.29 |
| Somatomotor | 307.26 | 703.17 | 41.76 | 0.44 | 0.810 | 0.14 | -0.51 | 0.79 |
| Default Mode | 208.18 | 373.64 | 41.13 | 0.56 | 0.953 | 0.18 | -0.48 | 0.84 |
| Lateral frontoparietal (right)<br>(control) | 1029.85 | 606.90 | 41.28 | 1.70 | 0.805 | 0.55 | -0.11 | 1.22 |
| Lateral frontoparietal (left)<br>(control) | 530.35 | 575.59 | 41.34 | 0.92 | 0.959 | 0.30 | -0.36 | 0.96 |
| T1-T3 Baseline - Follow-up (Patients) |  |  |  |  |  |  |  |  |
| Visual I | -1469.75 | 838.64 | 51.28 | -1.75 | 0.428 | -0.67 | -1.45 | 0.11 |
| Default mode (Frontomedial) | -1099.62 | 1095.75 | 51.54 | -1.00 | 0.936 | -0.38 | -1.15 | 0.39 |
| Motor | -827.68 | 1228.91 | 51.54 | -0.67 | 0.979 | -0.26 | -1.03 | 0.51 |
| Visual II | -2805.92 | 1136.71 | 48.99 | -2.47 | 0.137 | -0.97 | -1.78 | -0.16 |
| Salience | -103.95 | 427.18 | 50.61 | -0.24 | 0.973 | -0.09 | -0.87 | 0.68 |
| Visual III | -560.33 | 184.01 | 51.54 | -3.05 | 0.033* | -1.17 | -1.96 | -0.37 |
| Dorsolateral frontoparietal (attention) | 366.03 | 330.55 | 49.09 | 1.11 | 0.894 | 0.44 | -0.36 | 1.23 |
| Somatomotor | -957.12 | 828.98 | 51.46 | -1.15 | 0.810 | -0.44 | -1.21 | 0.33 |
| Default Mode | 163.77 | 449.05 | 49.04 | 0.36 | 0.953 | 0.14 | -0.65 | 0.93 |
| Lateral frontoparietal (right) (control) | 948.09 | 725.80 | 49.71 | 1.31 | 0.911 | 0.51 | -0.28 | 1.30 |
| Lateral frontoparietal (left) (control) | 318.27 | 686.96 | 49.98 | 0.46 | 0.959 | 0.18 | -0.60 | 0.96 |
| **T2-T3 Post Treatment - Follow-up (Patients)** |  |  |  |  |  |  |  |  |
| Visual I | -265.27 | 844.28 | 49.96 | -0.31 | 0.901 | -0.12 | -0.90 | 0.65 |
| Default mode (frontomedial) | -1174.82 | 1103.51 | 50.30 | -1.06 | 0.918 | -0.41 | -1.19 | 0.36 |
| Motor | -854.73 | 1237.61 | 50.30 | -0.69 | 0.979 | -0.27 | -1.04 | 0.51 |
| Visual II | -646.03 | 1139.52 | 46.96 | -0.57 | 0.936 | -0.22 | -1.02 | 0.57 |
| Salience | -253.10 | 429.61 | 49.08 | -0.59 | 0.973 | -0.23 | -1.01 | 0.55 |
| Visual III | -222.36 | 185.31 | 50.30 | -1.20 | 0.553 | -0.46 | -1.24 | 0.31 |
| Dorsolateral frontoparietal (attention) | 676.63 | 331.44 | 47.08 | 2.04 | 0.375 | 0.81 | 0.00 | 1.61 |
| Somatomotor | -1264.37 | 834.76 | 50.20 | -1.51 | 0.810 | -0.58 | -1.36 | 0.20 |
| Default Mode | -44.41 | 450.21 | 47.03 | -0.10 | 0.953 | -0.04 | -0.83 | 0.75 |
| Lateral frontoparietal (right)<br>(control) | -81.77 | 728.72 | 47.90 | -0.11 | 0.911 | -0.04 | -0.83 | 0.74 |
| Lateral frontoparietal (left)<br>(control) | -212.07 | 690.07 | 48.24 | -0.31 | 0.959 | -0.12 | -0.91 | 0.66 |
| **T1-T2 Baseline - Post (Healthy control group)** |  |  |  |  |  |  |  |  |
| Visual I | 2899.89 | 1164.33 | 54.59 | 2.49 | 0.142 | 1.33 | 0.23 | 2.42 |
| Default mode (frontomedial) | -1105.80 | 1519.62 | 54.64 | -0.73 | 0.936 | -0.39 | -1.45 | 0.68 |
| Motor | -397.51 | 1704.29 | 54.64 | -0.23 | 0.979 | -0.12 | -1.19 | 0.94 |
| Visual II | 1174.01 | 1596.51 | 53.65 | 0.74 | 0.936 | 0.41 | -0.70 | 1.52 |
| Salience | -718.41 | 594.88 | 54.40 | -1.21 | 0.973 | -0.65 | -1.74 | 0.43 |
| Visual III | 231.44 | 255.19 | 54.64 | 0.91 | 0.737 | 0.48 | -0.58 | 1.55 |
| Dorsolateral frontoparietal<br>(attention) | -60.20 | 464.02 | 53.71 | -0.13 | 0.897 | -0.07 | -1.18 | 1.04 |
| Somatomotor | 277.84 | 1150.03 | 54.63 | 0.24 | 0.810 | 0.13 | -0.94 | 1.19 |
| Default Mode | 1096.13 | 630.53 | 53.68 | 1.74 | 0.615 | 0.96 | -0.16 | 2.08 |
| Lateral frontoparietal (right)<br>(control) | 892.24 | 1015.38 | 54.04 | 0.88 | 0.911 | 0.48 | -0.62 | 1.58 |
| Lateral frontoparietal (left)<br>(control) | 482.45 | 959.68 | 54.16 | 0.50 | 0.959 | 0.27 | -0.82 | 1.36 |
| **Between subjects factors (Healthy - Patient)** |  |  |  |  |  |  |  |  |
| Visual I | 3022.45 | 977.23 | 62.60 | 3.09 | 0.030* | 1.38 | 0.45 | 2.31 |
| Default mode (frontomedial) | -1684.68 | 1268.67 | 62.78 | -1.33 | 0.860 | -0.59 | -1.48 | 0.31 |
| Motor | 1959.27 | 1422.84 | 62.78 | 1.38 | 0.839 | 0.61 | -0.28 | 1.51 |
| Visual II | 3446.14 | 1413.61 | 60.40 | 2.44 | 0.142 | 1.19 | 0.19 | 2.20 |
| Saliency | -632.19 | 506.44 | 62.06 | -1.25 | 0.973 | -0.57 | -1.50 | 0.35 |
| Visual III | 763.48 | 213.04 | 62.78 | 3.58 | 0.007** | 1.59 | 0.65 | 2.52 |
| Dorsolateral frontoparietal (attention) | 484.44 | 409.86 | 60.51 | 1.18 | 0.894 | 0.58 | -0.41 | 1.56 |
| Somatomotor | 650.07 | 961.64 | 62.73 | 0.68 | 0.810 | 0.30 | -0.59 | 1.19 |
| Default mode | 1062.92 | 557.58 | 60.46 | 1.91 | 0.430 | 0.93 | -0.06 | 1.93 |
| Lateral frontoparietal (right) (control) | -251.33 | 882.82 | 61.19 | -0.28 | 0.911 | -0.14 | -1.09 | 0.82 |
| Lateral frontoparietal (left) (control) | 525.13 | 829.05 | 61.46 | 0.63 | 0.959 | 0.30 | -0.65 | 1.24 |

**Table S5:**
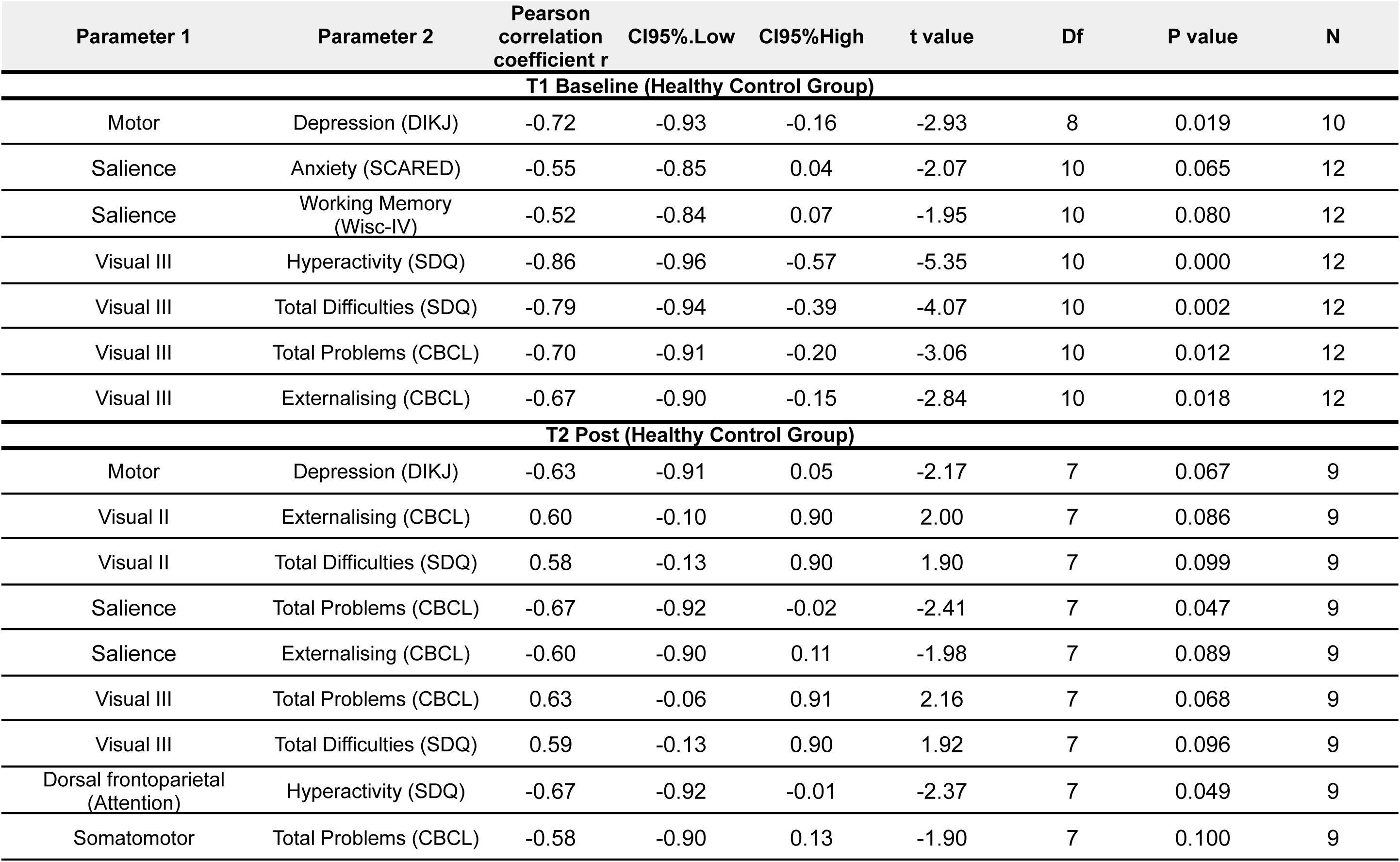
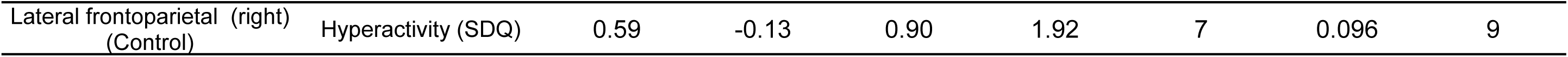
Significant Brain-Behaviour Correlations in the Healthy Control Group. Pearson correlation coefficients detailing the relationships between resting-state network activation and clinical/behavioural metrics in the Healthy Control Group. Correlations with unadjusted P Values (< 0.1) were selected and presented for distinct study timepoints (T1, T2)

**Table S6:**
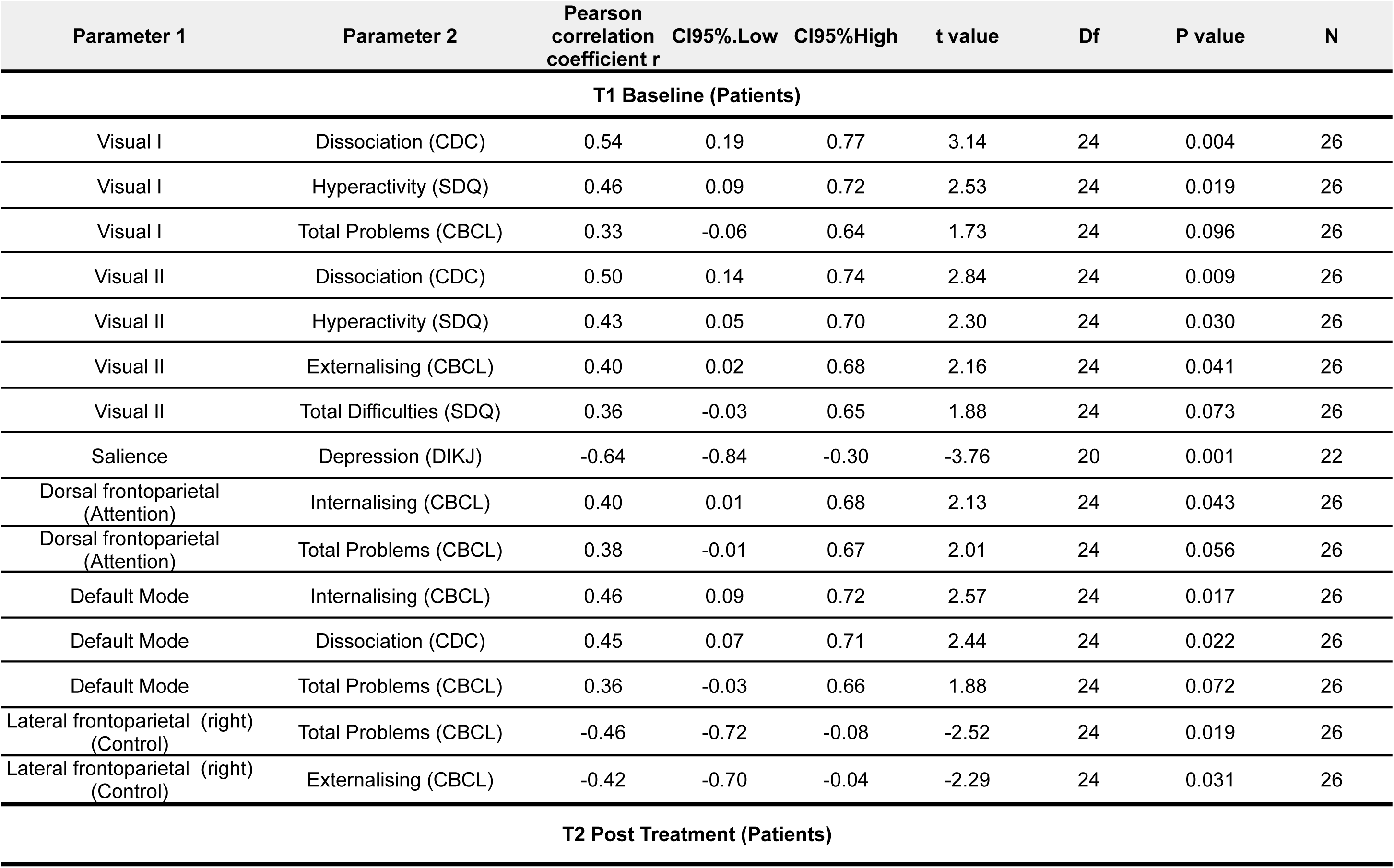
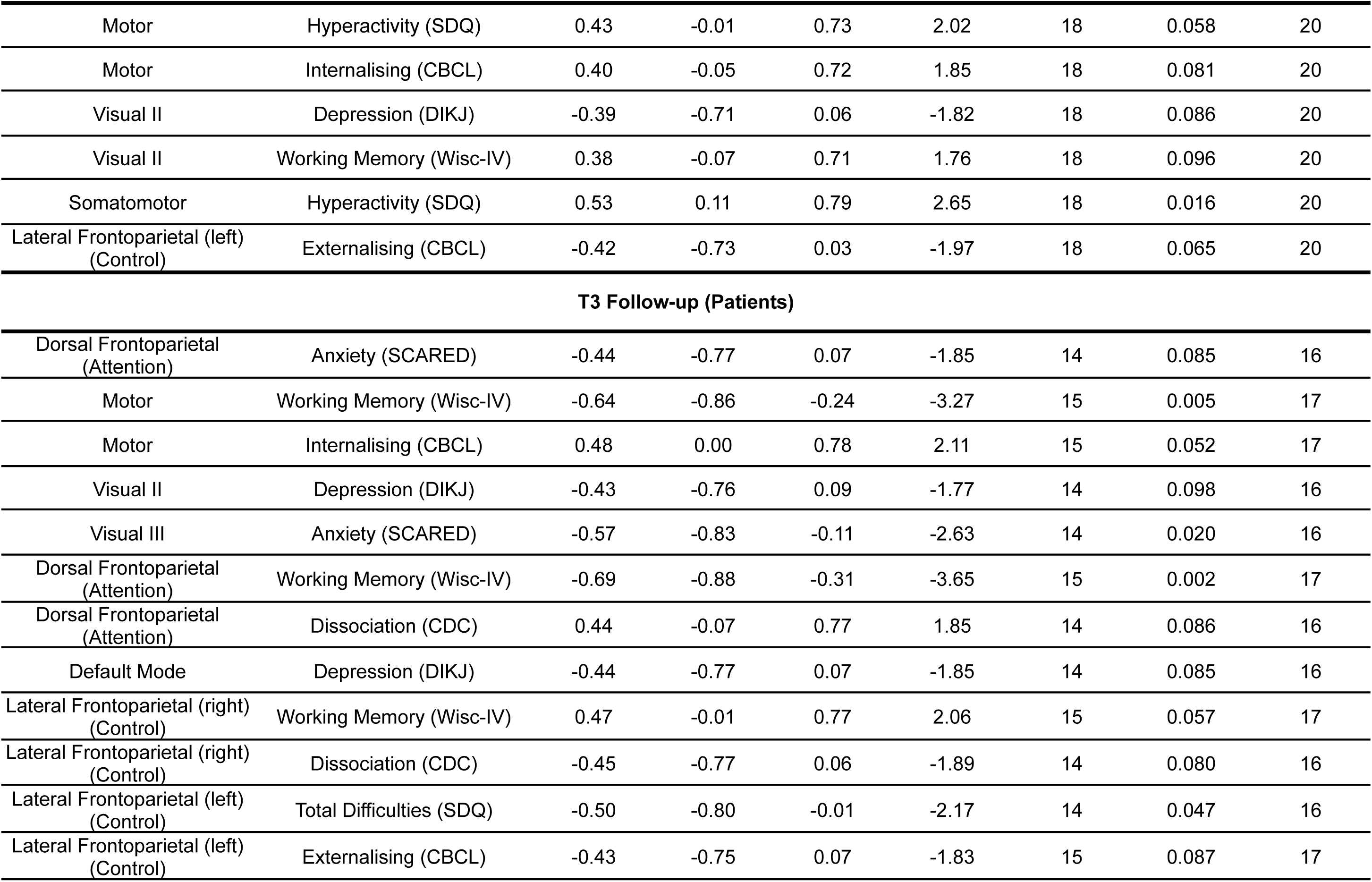
Significant Brain-Behaviour Correlations in the Intervention Group (Patients). Pearson correlation coefficients evaluating the longitudinal relationships between resting-state network activation and clinical symptom severity within the Intervention Group. Correlations with unadjusted P Values were selected and presented for distinct study timepoints (T1, T2, T3), to demonstrate that correlation patterns for visual networks and primary clinical outcome measures (eg. total problems and externalizing symptoms (CBCL) and hyperactivity (SDQ)) resolve even with higher threshold (p < 0.1). Abbreviations: **CBCL**, Child Behaviour Checklist; **SDQ**, Strengths and Difficulties Questionnaire; **WISC-IV,** Wechsler Intelligence Scale for Children, Fourth Edition; **PROPS**, Parent Report of Post-traumatic Symptoms; **SCARED,** The Screen for Child Anxiety Related Emotional Disorders; **DIKJ**, Depression Inventory for Children and Adolescents; **CDC**, Child Dissociative Checklist.

**Table S7:**
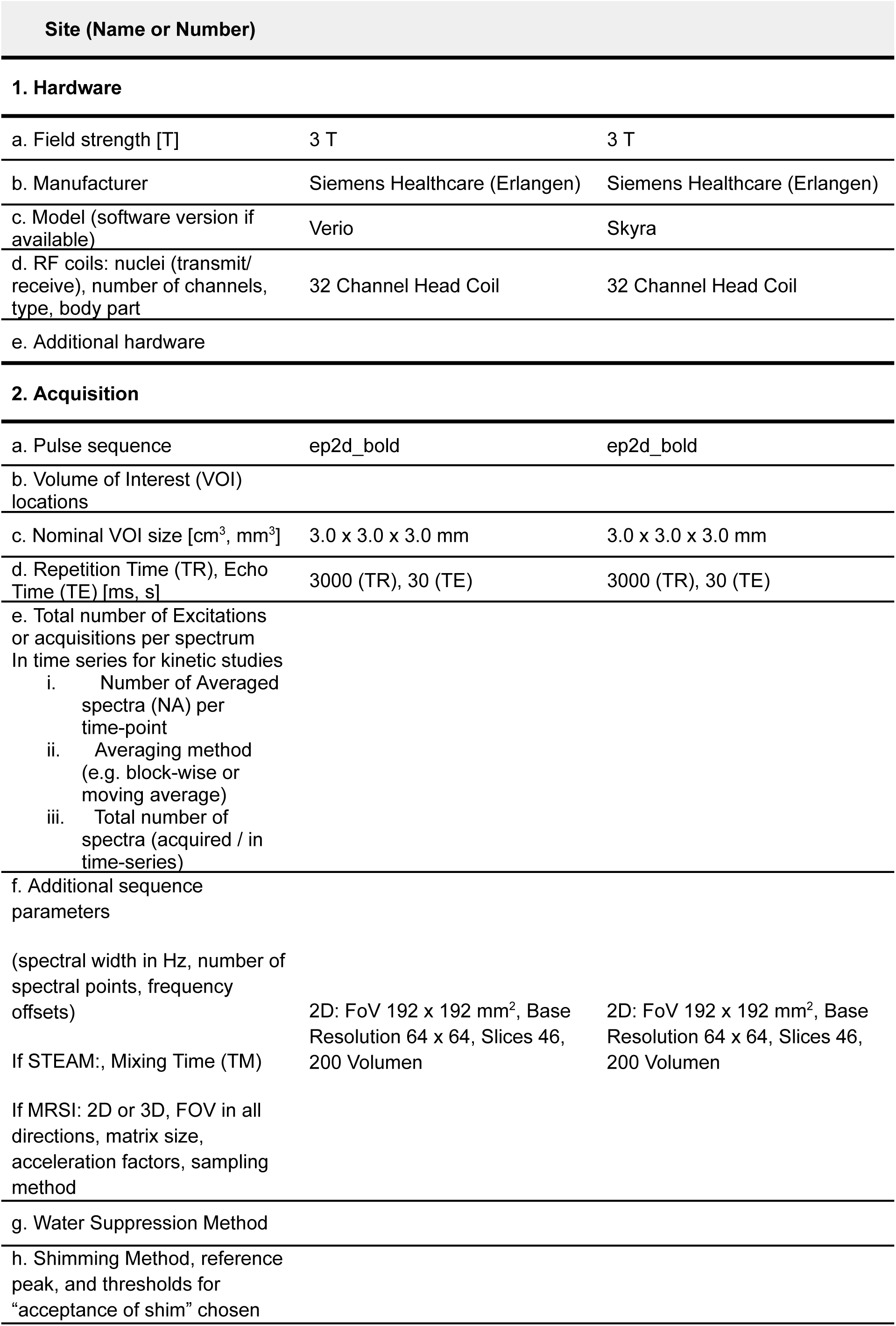
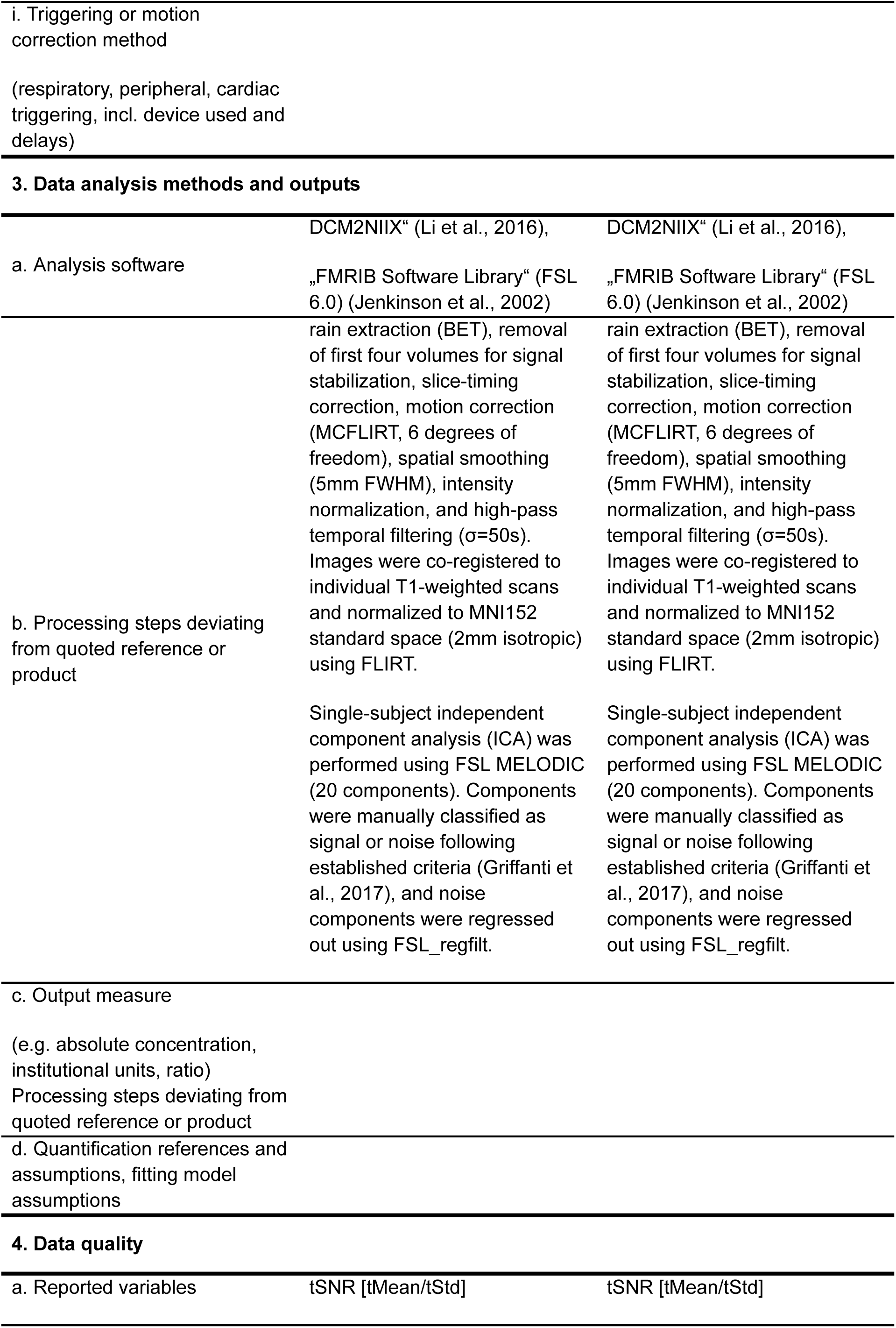
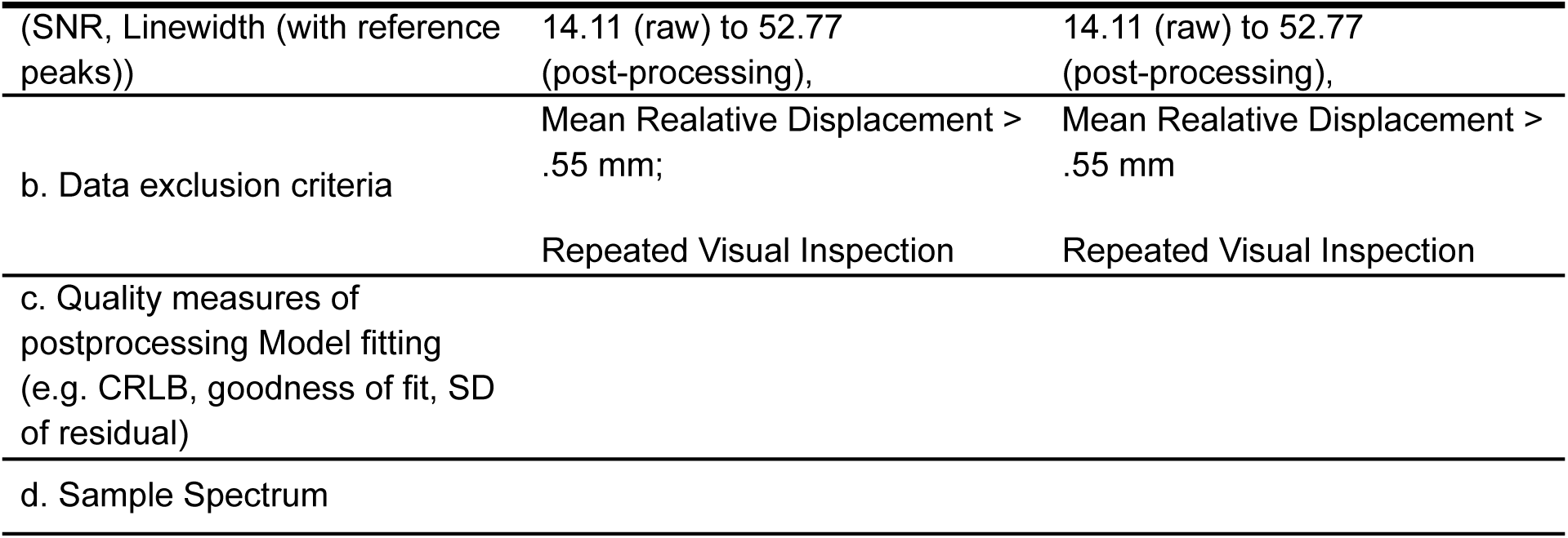
Standardized Neuroimaging Reporting and fMRI Parameters. Detailed specification of the MRI hardware, pulse sequences, acquisition parameters, data analysis software, and preprocessing pipelines utilized in the study

**Table S8:** Remission Rates on Clinical Variables. No dropouts were included in the analysis. Abbreviations: **CBCL**, Child Behaviour Checklist; **SDQ**, Strengths and Difficulties Questionnaire; **PROPS**, Parent Report of Post-traumatic Symptoms; **SCARED,** The Screen for Child Anxiety Related Emotional Disorders; **DIKJ**, Depression Inventory for Children and Adolescents; **CDC**, Child Dissociative Checklist. *Note: N (%). Participants excluded* from analysis because the symptom severity was not in the clinical range at T1 (baseline)*.

| Variable | T2 Post Treatment<br>(N=20) |  |  | T3 Follow-up<br>(N=14) |  |  |
| --- | --- | --- | --- | --- | --- | --- |
|  | Remission | Remained Clinical | Excluded* | Remission | Remained Clinical | Excluded* |
| **CBCL** |  |  |  |  |  |  |
| Total Problems | 9 (47.37%) | 10 (52.63%) | 1 | 6 (46.15%) | 7 (53.85%) | 1 |
| Externalizing | 11 (61.11%) | 7 (38.89%) | 2 | 7 (50.00%) | 7 (50.00%) | - |
| Internalizing | 5 (41.67%) | 7 (58.33%) | 8 | 5 (50.00%) | 5 (50.00%) | 4 |
| **SDQ** |  |  |  |  |  |  |
| Total Difficulties | 6 (42.86%) | 8 (57.14%) | 6 | 8 (66.67%) | 4 (33.33%) | 2 |
| Hyperactivity | 5 (41.67%) | 7 (58.33%) | 8 | 3 (33.33%) | 6 (66.67%) | 5 |
| **Trauma (PROPS)** | 11 (64.71%) | 6 (35.29%) | 3 | 8 (61.54%) | 5 (38.46%) | 1 |
| **Dissociation (CDC)** | 2 (28.57%) | 5 (71.43%) | 13 | 5 (71.43%) | 2 (28.57%) | 7 |
| **Anxiety (SCARED)** | 1 (33.33%) | 2 (66.67%) | 17 | 1 (100.00%) | - | 13 |
| **Depression (DIKJ)** | 5 (62.50%) | 3 (37.50%) | 12 | 5 (100.00%) | - | 9 |

**Figure S1:**
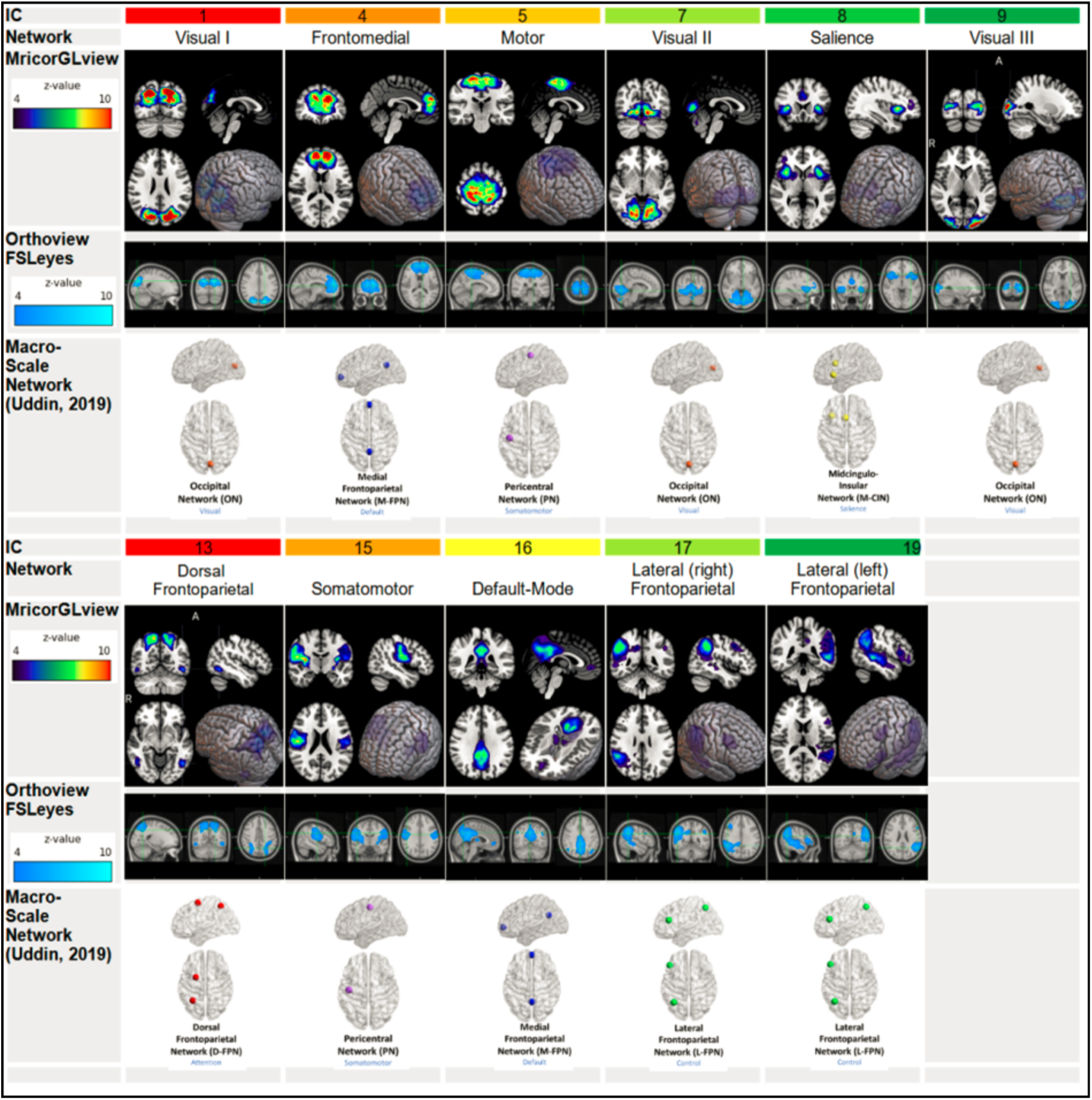
Spatial Topography of Identified Resting-State Functional Networks. Visualization of the functional resting-state networks extracted via grouped independent component analysis (group-ICA), displayed in radiological convention. Brain maps highlight z-score ranges between 4 and 10. The extracted independent components are structurally and functionally cross-referenced with the macro-scale resting-state network taxonomy proposed by Uddin et al. (2019). For group-ICAs, measurements with excessive noise (mean relative displacement > .55mm) and insufficient signal were excluded. Radiological convention shown on MRIcroGL and FSLeyes orthoview.

